# Independent Versus Joint Effects of Polygenic or Family-Based Schizophrenia Risk in Diverse Ancestry Youth in the ABCD Study

**DOI:** 10.1101/2025.05.16.25327647

**Authors:** Mahnoor Hyat, Jinhan Zhu, Toni A. Boltz, Matthew P. Conomos, Dylan E. Hughes, Alison E. Fohner, Katherine T. Foster, Tim B. Bigdeli, Jennifer K. Forsyth

## Abstract

**Background:** Subtle behavioral and cognitive symptoms often precede schizophrenia (SCZ) onset and are found in individuals with elevated risk for SCZ based on family history (SCZ-FH) and polygenic risk scores (SCZ-PRS). However, most SCZ-PRS studies focus on European ancestry youth, limiting generalizability. Furthermore, it remains unclear whether SCZ-FH reflects common variant polygenic risk or broader SCZ liability.

**Method:** Using baseline data from the Adolescent Brain Cognitive Development (ABCD) study, we investigated associations of a continuous SCZ-FH measure and SCZ-PRS, derived using PRS-CSx, with cognitive, behavioral and emotional measures derived from the NIH-Toolbox, Child Behavior Checklist (CBCL), and Kiddie Schedule for Affective Disorders and Schizophrenia (KSADS) for 9,636 children (mean age=9.92 yrs, 47.4% female), specifically 5,636 European, 2,093 African, and 1,477 Admixed American ancestry individuals.

**Results:** SCZ-FH was associated with elevated SCZ-PRS (b=0.39, FDR-p=0.02) and subthreshold psychotic symptom severity (b=3.74, FDR-p=0.01) in European youth, higher CBCL scores across ancestries (b range=2.90-4.81, FDR-p<0.001), and higher odds of depression, anxiety, conduct disorder, post-traumatic stress disorder, and attention-deficit/hyperactivity disorder (OR=2.17-5.09, FDR-p<0.001) across ancestries. SCZ-PRS was associated with lower cognitive functioning across ancestries (b=-0.43, FDR-p=0.023), CBCL total problems, anxious/depressed, rule-breaking and aggressive behaviors in European youth (b range=0.16-0.33, FDR-p<0.04), and depressive disorders in Admixed American youth (OR=1.37, FDR-p=0.02). Results remained consistent when SCZ-PRS and SCZ-FH were jointly modeled.

**Conclusion:** SCZ-FH showed stronger associations with broad psychopathology, while SCZ-PRS was associated with cognitive functioning and specific emotional and behavioral symptoms in European youth. These findings highlight their complementary role in early SCZ risk assessment and underscore the need to improve PRS utility across ancestries.

## Introduction

Schizophrenia (SCZ) is a severe psychiatric disorder with significant personal and societal impact (Chong et al., 2016; Evensen et al., 2016; Kar & Jain, 2016; Laursen, Nordentoft & Mortensen, 2014). Standard treatments typically begin after the onset of full-blown psychosis but are often insufficient in restoring premorbid levels of functioning. The clinical staging model of SCZ posits that persistent illness is a late-stage manifestation of disrupted neurodevelopmental processes and that subtle signs and symptoms may be evident early on that could serve as markers of future psychopathology (Forsyth & Lewis, 2017; McGorry et al., 2014). For example, birth cohort studies have demonstrated that individuals who later receive diagnoses of SCZ have higher rates of cognitive deficits and behavioral and emotional problems in childhood (Mollon & Reichenberg, 2018; Riglin et al., 2017; Welham, Isohanni, Jones, & McGrath, 2009). Elevated psychotic-like experiences (PLEs) in childhood, or subthreshold psychotic symptoms, are also associated with later psychotic disorder onset (Healy et al., 2019; Laurens et al., 2007). Similarly, adults with SCZ are more likely to have met criteria for other psychiatric diagnoses, such as anxiety and affective disorders, in childhood and adolescence (Maibing et al., 2015). This suggests that childhood behavioral and clinical signs may help identify individuals at risk for SCZ; however, as most children with cognitive or emotional difficulties do not develop SCZ, a clearer understanding of how these symptoms relate to underlying etiological processes is essential.

Importantly, genetic factors are known to contribute to the development of SCZ, with twin studies estimating heritability at around 80% (Hilker et al., 2018). Given elevated rates of SCZ among individuals with a family history of psychosis (SCZ-FH) (Cheng et al., 2018; Goldstein, Buka, Seidman, & Tsuang, 2010), SCZ-FH has long served as a proxy for genetic risk and a means of studying potential antecedents of SCZ in youth at elevated risk (Díaz-Castro, Hoffman, Cabello-Rangel, Arredondo, & Herrera-Estrella, 2021; Niemi, Suvisaari, Tuulio-Henriksson, & Lönnqvist, 2003). For example, offspring of individuals with SCZ have been shown to exhibit higher rates of lifetime psychiatric disorders and cognitive impairments (Erlenmeyer-Kimling et al., 2000; Keshavan, 2009; Sanchez-Gistau et al., 2015). However, the extent to which SCZ-FH captures genetic-specific versus broader risk factors for SCZ is unclear. Furthermore, many individuals who develop SCZ do not have an immediate family member with the disorder, raising questions about the generalizability of cognitive and clinical markers identified in youth with SCZ-FH to those who develop SCZ without known familial risk.

Fortunately, advancements in genetic analysis, especially the advent of large-scale genome-wide association studies (GWAS), have enabled the identification of robust associations between psychiatric disorders and specific genetic risk variants (Schizophrenia Working Group of the Psychiatric Genomics Consortium, 2014). The most recent GWAS for SCZ, based on 76,755 patients and 243,649 controls, identified 287 distinct loci that were significantly associated with SCZ (Trubetskoy et al., 2022). This allows for the computation of polygenic risk scores (PRS), which capture each individuals’ genetic risk for the disorder as a weighted sum of risk alleles (Agerbo et al., 2015; Hujoel, Loh, Neale, & Price, 2022; Legge et al., 2019), and can be tested for association with phenotypes in childhood to identify genetically-mediated antecedents of SCZ. SCZ-PRS, which can explain up to 7.3-7.7% of the variance in SCZ case-control status (Legge et al., 2019; Trubetskoy et al., 2022), have been associated with negative symptoms and cognitive impairments in childhood, but findings for depressive symptoms and psychotic experiences are mixed (Jones et al., 2016; Mistry, Harrison, Smith, Escott-Price, & Zammit, 2018; Nivard et al., 2017).

Beyond equivocal links between SCZ-PRS and childhood psychopathology, a key challenge remains in developing generalizable SCZ-PRS to improve prediction across populations. This challenge is due to an over-representation of European ancestry individuals in existing GWAS, which reduces PRS accuracy in diverse populations because of differences in allele frequency and linkage disequilibrium (LD) patterns (Kachuri et al., 2024). However, recent efforts to diversify GWAS samples (Bigdeli et al., 2020; Nguyen et al., 2022) and the development of advanced PRS construction methods like PRS-CSx, which refine allele effect size estimates by integrating multi-ancestry GWAS summary statistics and ancestry-matched LD panels (Ruan et al., 2022), are improving cross-population accuracy of PRS and supporting more equitable risk assessment.

To investigate whether associations between early cognitive and behavioral signs and symptoms and SCZ-FH or SCZ-PRS represent independent or overlapping aspects of SCZ risk, and to assess the generalizability of these associations across diverse genetic ancestries, the current study used data from the Adolescent Brain Cognitive Development (ABCD) Study to examine how SCZ-FH and SCZ-PRS relate to cognitive, behavioral, and emotional functioning during childhood. The ABCD Study is the largest nationally representative, longitudinal study of child brain development in the US and includes genetic, behavioral and clinical information.

Previous studies in ABCD found that higher SCZ-PRS were linked to worse cognitive functioning, greater attentional variability, and more psychotic-like experiences, but showed mixed results for internalizing and externalizing problems in childhood (Chang et al., 2024; Loughnan et al., 2022; Wainberg, Jacobs, Voineskos, & Tripathy, 2022). However, these studies restricted primary analyses to European ancestry youth and/or used methods for PRS construction that were not optimized for cross-ancestry PRS accuracy. By incorporating SCZ GWAS summary statistics from ancestrally diverse populations and using PRS-CSx to optimize SCZ-PRS accuracy across groups, the current study aims to refine prior investigations for diverse youth and determine whether integrating SCZ-FH and SCZ-PRS into a combined model yields incremental utility for identifying and differentiating early SCZ risk.

## Methods and Materials

### Sample

This study utilized data from the ABCD study which enrolled 11,880 children across 21 sites to reflect the sociodemographic characteristics of the general US population. Using data from Annual Data Release 4.0 (Yang & Jernigan, Terry, n.d.), we focused on baseline assessment data when children were 9-10 years old to investigate SCZ-related signs and symptoms at the earliest time possible in the ABCD study. Analyses were conducted for youth of European American (EUR; *n* = 5,626), African American (AFR; *n* = 2,093), or Admixed American (AMR; *n* = 1,477) ancestry (47.4% female; mean age=9.92 years), as determined by genetic similarity to HapMap3 reference populations, as these groups were sufficiently powered for investigation.

### Measures

#### SCZ Risk

SCZ-FH was assessed by the ABCD team using the caregiver-reported Family History Assessment Survey which collected information on family members’ experiences with depression, mania, hallucinations, and related conditions. SCZ-FH was determined through the question: *“Has ANY blood relative of your child ever had a period lasting six months when they saw visions or heard voices or thought people were spying on them or plotting against them?”* A continuous weighted endorsement measure was created as part of the current project by summing the number of affected relatives weighted by the degree of relatedness for each affected relative (0.5 for first-degree and 0.25 for second-degree relatives).

The second SCZ risk measure was a molecularly defined SCZ-PRS. These scores were derived from the imputed genotyping data that underwent rigorous quality control by the ABCD team following the Ricopili pipeline (Lam et al., 2020; Wainberg et al., 2022). Imputation was completed using the Trans-Omics for Precision Medicine (TOPMed) reference panel (Das et al., 2016; Loh et al., 2016; Taliun et al., 2021). Following ABCD recommendations, we excluded data from plate 461 due to poor quality. Only high-quality imputed variants (i.e., R2 > 0.8) with a minor allele frequency (MAF) > 0.01 were retained for PRS construction. To minimize confounding caused by the complex LD structure of the major histocompatibility complex (MHC) region, only one SNP in the region with the strongest association to SCZ was retained (Loughnan et al., 2022; Trubetskoy et al., 2022).

Before deriving SCZ-PRS, PRS-CSx was employed to refine the effect size estimates for each SNP by integrating summary statistics from multiple GWAS and accounting for LD patterns across populations. This method was chosen due to its demonstrated accuracy in improving cross-ancestry PRS accuracy compared to many other PRS construction methods (Ruan et al., 2022). Specifically, summary statistics were obtained from the largest GWAS of SCZ, in which European ancestry individuals were over-represented (Trubetskoy et al., 2022), as well as smaller GWAS focused on Admixed American and African ancestry individuals (Bigdeli et al., 2021).

The PRS-CSx output included a meta-analysis file, which provided refined SNP effect sizes across populations using an inverse-variance-weighted meta-analysis of population-specific posterior effect size estimates after incorporating 1000 Genomes Project LD reference panels. It also provided ancestry-specific SNP effect size estimates for each GWAS. The PRS-CSx meta-analyzed SNP estimates were used to construct SCZ-PRS for Admixed American and African ancestry individuals, whereas, for the European ancestry group, we used SNP effect size estimates derived from the predominantly European ancestry GWAS as prior evidence suggests meta-analyzed effect size estimates improve prediction for non-European groups, whereas ancestry-specific estimates offer greater accuracy for European populations (Ruan et al., 2022). SCZ-PRS were then generated in PLINK before being z-score standardized within each ancestry group.

#### Behavioral and Cognitive Signs and Symptoms

##### I) Dimensional Assessments

Children completed tasks from the NIH Cognitive Toolbox (NIH-TB) which quantifies cognitive functioning across domains such as working memory, language and inhibitory control (Gershon et al., 2013; Weintraub et al., 2013). A winsorized, age-corrected composite cognition score, summarizing performance across tasks, was used for analysis. Primary caregivers also completed the Child Behavior Checklist (CBCL) to provide a measure of emotional and behavioral problems in children over the past 6 months (Achenbach, 1991). The CBCL total composite score, along with scores for the eight primary subscales: anxious/depressed, withdrawn/depressed, somatic complaints, social problems, thought problems, attention problems, rule-breaking behavior, and aggressive behavior severity were used for analysis.

Additionally, children completed the Prodromal Questionnaire – Brief Child Version (PQ-BC) which is a 21-item scale that assesses occurrences of psychotic-like experiences and level of distress for any endorsed experience. PLEs were operationalized as the sum of the number of endorsed items weighted by their associated distress (PQ-BC Distress) (Chang et al., 2024).

##### II) Diagnostic Assessments

Caregivers and youth completed the semi-structured, self-administered computerized Kiddie Schedule for Affective Disorders and Schizophrenia (KSADS-COMP), which assesses mental health conditions, including depression, anxiety disorders, conduct disorder and ADHD (Kaufman et al., 1997). Lifetime history of diagnoses were derived from the caregiver self-administered KSADS-COMP, as they were shown to have greater concordance with gold-standard clinician interviews integrating parent and youth report, compared to diagnoses derived from the youth self-administered KSADS-COMP (Townsend et al., 2020). To facilitate comparison with alternate methods for deriving ADHD diagnoses, we generated an additional ADHD variable using CBCL cut-offs (Cordova et al., 2022). A T-score of ≥65 on the CBCL Attention Problems scale was established as the threshold for clinical-level attention problems. See Supplementary Methods for details on dimensional and diagnostic assessments and *n*s available per measure.

### Statistical Analysis

#### Ancestry Principal Components, Ancestry Grouping, and Genetic Relatedness

To create ancestrally homogeneous groups of subjects, we conducted principal components analysis (PCA) using high-quality SNPs from the ABCD dataset merged with HapMap3 data as the reference (The International HapMap 3 Consortium, 2010). Ancestry representative PCs were generated following an iterative procedure optimized for samples from diverse ancestral groups with familial relatedness (Conomos, Laurie, et al., 2016). This involved using PC-AiR for PC calculation and PC-Relate for kinship coefficient estimation, implemented in the GENESIS R package (Conomos, Miller, & Thornton, 2015; Conomos, Reiner, Weir, & Thornton, 2016). We utilized the top eight PCs as inputs for a Random Forest classifier to categorize participants into ancestry groups based on genetic similarity, using HapMap3 super-population labels (Alexander & Lange, 2011). See Figure S1 for distributions across the PCs.

#### Association Testing

We employed linear and logistic mixed models, implemented using the GENESIS package in R (Gogarten et al., 2019) to examine associations. Associations between each phenotype and either SCZ-FH or SCZ-PRS, were first assessed in independent models (i.e. dependent variable ∼ SCZ-FH/PRS + covariates), followed by a joint model including both SCZ-FH and SCZ-PRS (i.e., dependent variable ∼ SCZ-FH + SCZ-PRS + covariates) to refine coefficient estimates for each SCZ risk measure, while accounting for the effect of the other. We conducted analyses separately for the three ancestries (EUR-only, AFR-only and AMR-only), followed by a combined analysis using within-ancestry z-scored SCZ-PRS (cross-ancestry). The fixed-effect covariates included sex, age, study site and ancestry principal components, while genetic correlation was accounted for as random effect through a genetic relatedness matrix (GRM) generated using PC-Relate. P-values for all dimensional measures models were corrected for multiple testing using false discovery rate (FDR) correction for the full sample and within each ancestry, across both risk SCZ measures (i.e. *n* models = 20 per group). P-values for all diagnostic outcomes were similarly FDR-corrected for multiple testing for the full sample and within each ancestry, across both risk SCZ measures (i.e. *n* models = 12 per group). The proportions of variance explained (PVE) by each risk factor, both independently and jointly, were estimated using score statistics (Hu et al., 2021). We also conducted a sensitivity analysis incorporating income-to-needs ratio as an additional covariate to determine whether poverty-related factors influence SCZ-FH & SCZ-PRS associations with psychopathology. See Supplementary Methods for information on how this covariate was derived.

## Results

We first assessed associations between SCZ-FH and SCZ-PRS; as expected, they showed a positive relationship across ancestries (Figure S2). This association was significant for the EUR-only group (*b*=0.39, p=0.01, FDR p=0.02), but not in the cross-ancestry analysis (*b=*0.096, p=0.27, FDR p=0.53), or for AFR-only (*b=*0.07, p=0.53, FDR p=0.53) and AMR-only models (*b=*0.14, p=0.51, FDR p=0.53).

### Independent Associations Between SCZ Risk Measures & Dimensional Measures

Table 1 and Figure S3 summarize the independent associations between SCZ-FH and SCZ-PRS and potential childhood dimensional phenotypes related to SCZ. For NIH-TB total cognition, no significant associations were observed with SCZ-FH in the cross-ancestry analysis, nor in the EUR-only, AFR-only and AMR-only groups. Conversely, greater SCZ-FH was associated with higher CBCL total problems scores in the cross-ancestry group (*b*=9.04, p<0.001, FDR p<0.001) and within each of the three ancestry groups (*b* range=7.53-10.16, p<=0.002, FDR p<0.001). SCZ-FH also showed robust associations with all eight CBCL subscales in the cross-ancestry analysis and within each of the three groups (*b* range=2.90-4.81, p<0.001, FDR p<0.001). Additionally, individuals with greater SCZ-FH had higher PQB distress scores in the cross-ancestry group (*b*=3.45, p<0.001, FDR p<0.001) and in EUR-only (*b*=3.74, p=0.003, FDR p=0.01), although this association was only nominally significant in the AMR-only group and null in the AFR-only group.

**Table 1.**
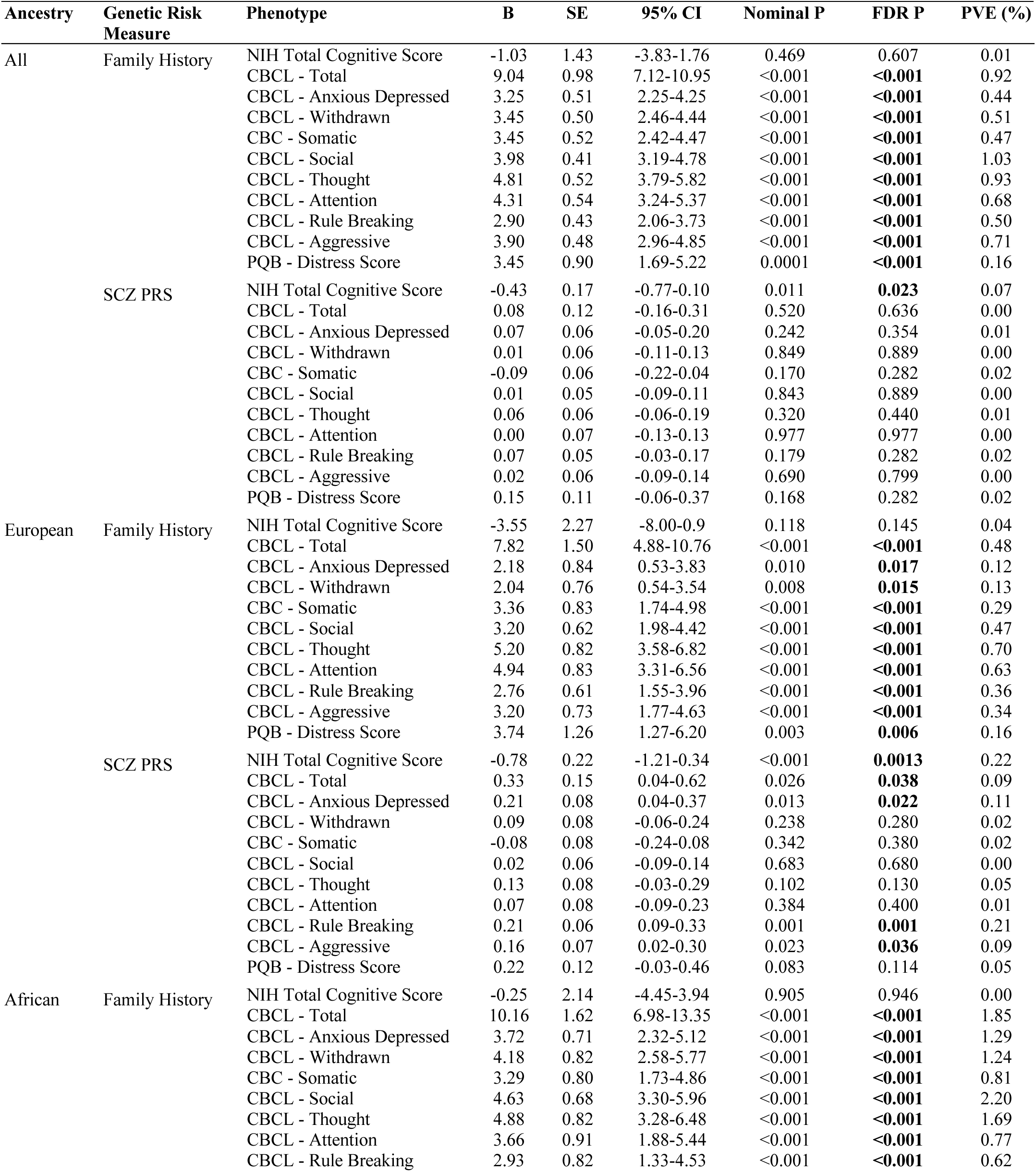
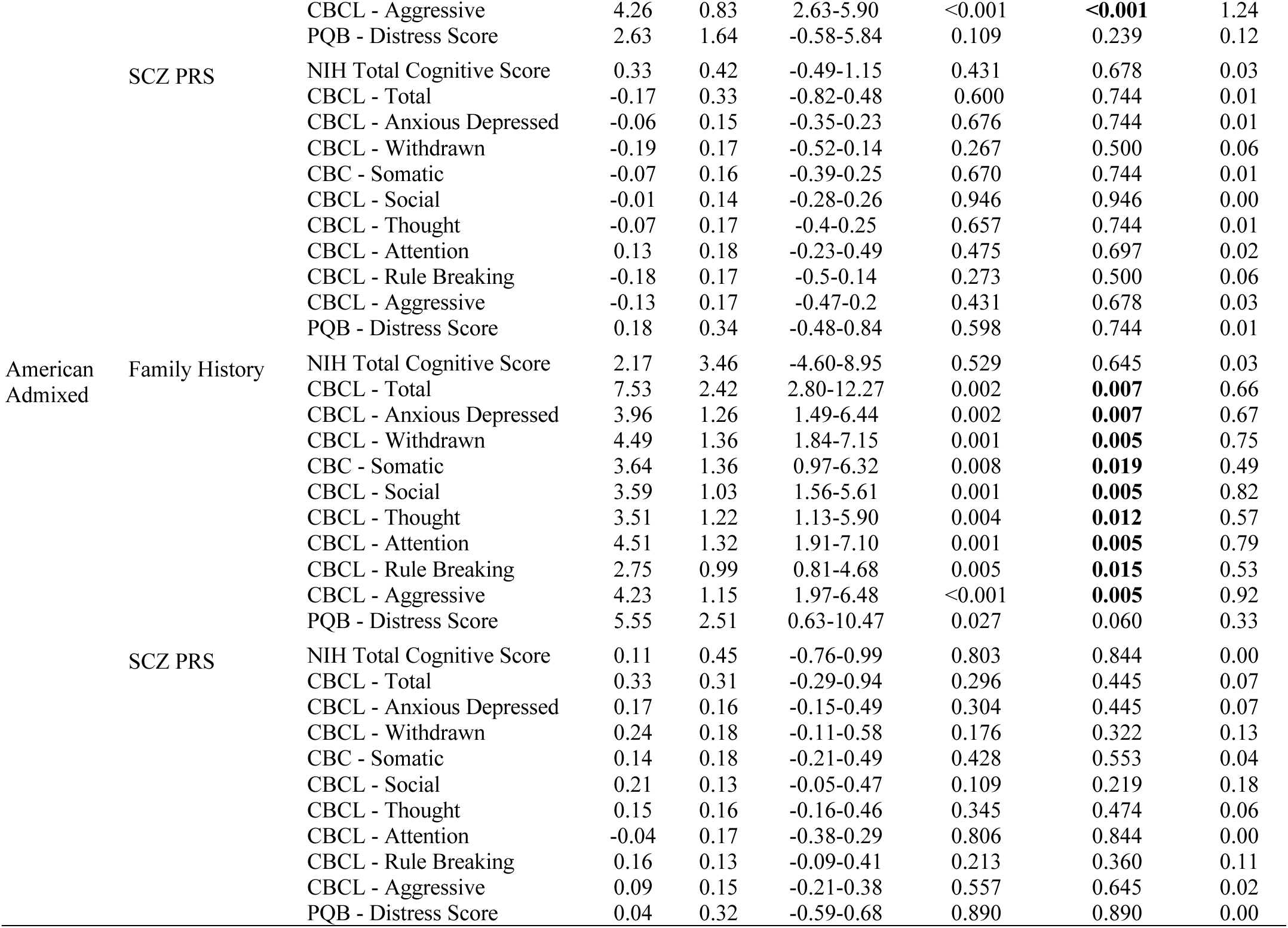
Independent associations of schizophrenia polygenic risk scores (SCZ-PRS) and family history of psychosis (SCZ-FH) with dimensional, total cognitive score derived from the NIH-Toolbox, Child Behavior Checklist (CBCL) scores, and prodromal questionnaire (PQB) scores. FDR P refers to the false discovery rate–corrected p value; PVE refers to the percentage of variance explained.

Higher SCZ-PRS was associated with significantly lower NIH-TB total cognition scores in the cross-ancestry analysis (*b*=-0.43, p=0.01 FDR p=0.02), as well as within EUR-only (*b*= −0.78, p<0.001, FDR p=0.001), but not in the AFR-only or AMR-only groups. SCZ-PRS was not significantly associated with CBCL total problems score (*b*=0.08 p=0.52, FDR p=0.64) or scores for any of the 8 subscales in the cross-ancestry analysis (*b* range=-0.09-0.07, p=0.17-0.98, FDR p=0.28-0.98). However, within the EUR-only group, higher SCZ-PRS was significantly associated with higher CBCL total problems (*b*=0.33, p=0.03, FDR p=0.04), anxious/depressed symptoms (*b*=0.21, p=0.01, FDR p=0.02), rule-breaking (*b*=0.21, p=0.001, FDR p=0.001) and aggressive behaviors (*b*=0.16, p=0.02, FDR p=0.04). SCZ-PRS and PQB Distress scores were not associated in the cross-ancestry analysis (*b*=0.15, p=0.17, FDR p=0.28) or within any of the three ancestries (*p*s>.05).

### Independent associations between SCZ Risk Measures & Diagnoses in Childhood

Table 2 and Figure S4 show the independent associations between the SCZ risk measures and clinical diagnoses in childhood. SCZ-FH was significantly associated with lifetime history of multiple diagnoses. In the cross-ancestry analysis, SCZ-FH was associated with higher likelihood of a depressive disorder (*OR*=2.95 p<0.001, FDR p<0.001), conduct disorder (*OR*=2.18, p<0.001, FDR p<0.001), anxiety disorder excluding (*OR*=4.75, p<0.001, FDR p<0.001), or including PTSD (*OR*=5.09, p<0.001, FDR p<0.001), and ADHD (*OR*=2.73, p<0.001, FDR p<0.001). The alternate CBCL-based ADHD diagnosis history was also associated with SCZ-FH (*OR*=4.41, p<0.001, FDR p<0.001). Associations within specific ancestries were similar, including showing significant associations across diagnoses for AFR-only; however, the associations for depressive and conduct disorders were not significant in EUR-only, and the association for depressive disorders was not significant in AMR-only. Additionally, the association for conduct disorder in AMR-only youth did not survive multiple testing correction.

**Table 2.** Independent associations of schizophrenia polygenic risk scores (SCZ-PRS) and family history of psychosis (SCZ-FH) with diagnostic phenotypes derived from the Kiddie Schedule for Affective Disorders and Schizophrenia (KSADS) or Child Behavior Checklist (CBCL). FDR P refers to the false discovery rate–corrected p value; PVE refers to the percentage of variance explained.

| Ancestry | Genetic Risk Measure | Phenotype | OR | SE | 95% CI | Nominal P | FDR P | PVE (%) |
| --- | --- | --- | --- | --- | --- | --- | --- | --- |
| All | Family History | KSADS - ADHD | 2.73 | 0.19 | 1.87-3.98 | <0.001 | **<0.001** | 0.35 |
|  |  | CBCL - ADHD | 4.41 | 0.23 | 2.78-6.97 | <0.001 | **<0.001** | 0.63 |
|  |  | KSADS - Anxiety Disorders | 4.75 | 0.20 | 3.20-7.05 | <0.001 | **<0.001** | 0.89 |
|  |  | KSADS - Anxiety Disorders w/ PTSD | 5.09 | 0.20 | 3.45-7.52 | <0.001 | **<0.001** | 0.99 |
|  |  | KSADS - Depressive Disorders | 2.95 | 0.27 | 1.75-4.97 | <0.001 | **<0.001** | 0.24 |
|  |  | KSADS - Conduct Disorder | 2.18 | 0.21 | 1.44-3.30 | <0.001 | **<0.001** | 0.18 |
|  | SCZ PRS | KSADS - ADHD | 1.03 | 0.03 | 0.98-1.08 | 0.293 | 0.440 | 0.01 |
|  |  | CBCL - ADHD | 0.96 | 0.04 | 0.88-1.04 | 0.277 | 0.440 | 0.02 |
|  |  | KSADS - Anxiety Disorders | 1.03 | 0.03 | 0.97-1.09 | 0.356 | 0.455 | 0.01 |
|  |  | KSADS - Anxiety Disorders w/ PTSD | 1.03 | 0.03 | 0.97-1.09 | 0.379 | 0.455 | 0.01 |
|  |  | KSADS - Depressive Disorders | 1.03 | 0.05 | 0.94-1.13 | 0.486 | 0.530 | 0.01 |
|  |  | KSADS - Conduct Disorder | 1.01 | 0.03 | 0.95-1.07 | 0.707 | 0.707 | 0.00 |
| European | Family History | KSADS - ADHD | 2.81 | 0.32 | 1.51-5.22 | 0.001 | **0.003** | 0.23 |
|  |  | CBCL - ADHD | 6.69 | 0.40 | 3.07-14.58 | <0.001 | **<0.001** | 0.57 |
|  |  | KSADS - Anxiety Disorders | 4.03 | 0.32 | 2.15-7.54 | <0.001 | **<0.001** | 0.43 |
|  |  | KSADS - Anxiety Disorders w/ PTSD | 4.06 | 0.32 | 2.18-7.57 | <0.001 | **<0.001** | 0.44 |
|  |  | KSADS - Depressive Disorders | 2.89 | 0.54 | 1.00-8.33 | 0.050 | 0.100 | 0.09 |
|  |  | KSADS - Conduct Disorder | 1.41 | 0.36 | 0.69-2.86 | 0.343 | 0.374 | 0.02 |
|  | SCZ PRS | KSADS - ADHD | 1.08 | 0.03 | 1.01-1.16 | 0.024 | 0.058 | 1.05 |
|  |  | CBCL - ADHD | 1.02 | 0.06 | 0.91-1.14 | 0.720 | 0.720 | 0.00 |
|  |  | KSADS - Anxiety Disorders | 1.06 | 0.04 | 0.99-1.14 | 0.118 | 0.142 | 0.05 |
|  |  | KSADS - Anxiety Disorders w/ PTSD | 1.07 | 0.04 | 0.99-1.15 | 0.079 | 0.130 | 0.07 |
|  |  | KSADS - Depressive Disorders | 1.11 | 0.07 | 0.97-1.27 | 0.115 | 0.142 | 0.06 |
|  |  | KSADS - Conduct Disorder | 1.07 | 0.04 | 0.99-1.15 | 0.086 | 0.130 | 0.06 |
| African | Family History | KSADS - ADHD | 2.34 | 0.29 | 1.33-4.12 | 0.003 | **0.006** | 0.50 |
|  |  | CBCL - ADHD | 3.06 | 0.33 | 1.60-5.88 | 0.001 | **0.003** | 0.78 |
|  |  | KSADS - Anxiety Disorders | 5.12 | 0.31 | 2.81-9.33 | <0.001 | **<0.001** | 2.05 |
|  |  | KSADS - Anxiety Disorders w/ PTSD | 6.13 | 0.30 | 3.40-11.08 | <0.001 | **<0.001** | 2.57 |
|  |  | KSADS - Depressive Disorders | 3.18 | 0.35 | 1.60-6.33 | 0.001 | **0.003** | 0.74 |
|  |  | KSADS - Conduct Disorder | 2.66 | 0.30 | 1.47-4.81 | 0.001 | **0.003** | 0.64 |
|  | SCZ PRS | KSADS - ADHD | 1.01 | 0.07 | 0.89-1.15 | 0.873 | 0.932 | 0.00 |
|  |  | CBCL - ADHD | 0.98 | 0.09 | 0.82-1.17 | 0.812 | 0.932 | 0.00 |
|  |  | KSADS - Anxiety Disorders | 1.03 | 0.08 | 0.88-1.21 | 0.698 | 0.932 | 0.01 |
|  |  | KSADS - Anxiety Disorders w/ PTSD | 1.01 | 0.08 | 0.86-1.17 | 0.932 | 0.932 | 0.00 |
|  |  | KSADS - Depressive Disorders | 0.89 | 0.10 | 0.74-1.08 | 0.245 | 0.420 | 0.08 |
|  |  | KSADS - Conduct Disorder | 1.02 | 0.07 | 0.88-1.18 | 0.806 | 0.932 | 0.00 |
| American<br>Admixed | Family History | KSADS - ADHD | 4.16 | 0.48 | 1.61-10.73 | 0.003 | **0.010** | 0.72 |
|  |  | CBCL - ADHD | 5.95 | 0.60 | 1.84-19.25 | 0.003 | **0.010** | 0.82 |
|  |  | KSADS - Anxiety Disorders | 5.78 | 0.52 | 2.08-16.08 | 0.001 | **0.009** | 1.05 |
|  |  | KSADS - Anxiety Disorders w/ PTSD | 5.25 | 0.52 | 1.89-14.59 | 0.001 | **0.009** | 0.93 |
|  |  | KSADS - Depressive Disorders | 2.73 | 0.71 | 0.69-10.88 | 0.154 | 0.264 | 0.16 |
|  |  | KSADS - Conduct Disorder | 3.15 | 0.57 | 1.02-9.70 | 0.046 | 0.092 | 0.33 |
|  | SCZ PRS | KSADS - ADHD | 0.95 | 0.07 | 0.82-1.10 | 0.503 | 0.670 | 0.04 |
|  |  | CBCL - ADHD | 0.91 | 0.11 | 0.74-1.13 | 0.406 | 0.608 | 0.06 |
|  |  | KSADS - Anxiety Disorders | 1.04 | 0.091 | 0.87-1.24 | 0.657 | 0.717 | 0.02 |
|  |  | KSADS - Anxiety Disorders w/ PTSD | 1.01 | 0.09 | 0.85-1.21 | 0.873 | 0.873 | 0.00 |
|  |  | KSADS - Depressive Disorders | 1.37 | 0.12 | 1.08-1.74 | 0.008 | **0.020** | 0.53 |
|  |  | KSADS - Conduct Disorder | 0.96 | 0.10 | 0.79-1.15 | 0.643 | 0.717 | 0.02 |

SCZ-PRS was not significantly associated with lifetime history of the assessed diagnoses in the cross-ancestry analysis or within individual ancestry groups for most disorders. However, there was a significant association between SCZ-PRS and depressive disorders among AMR-only youth (*OR*=1.37, p=0.01, FDR p=0.02).

### Joint modeling of SCZ Risk Measures Versus Dimensional Measures

Table 3 and Figure 1A summarize associations between the two SCZ risk measures and dimensional phenotypes when modeled jointly. These joint model associations closely mirrored patterns observed in the independent models when each SCZ risk measure was tested separately.

**Figure 1.**
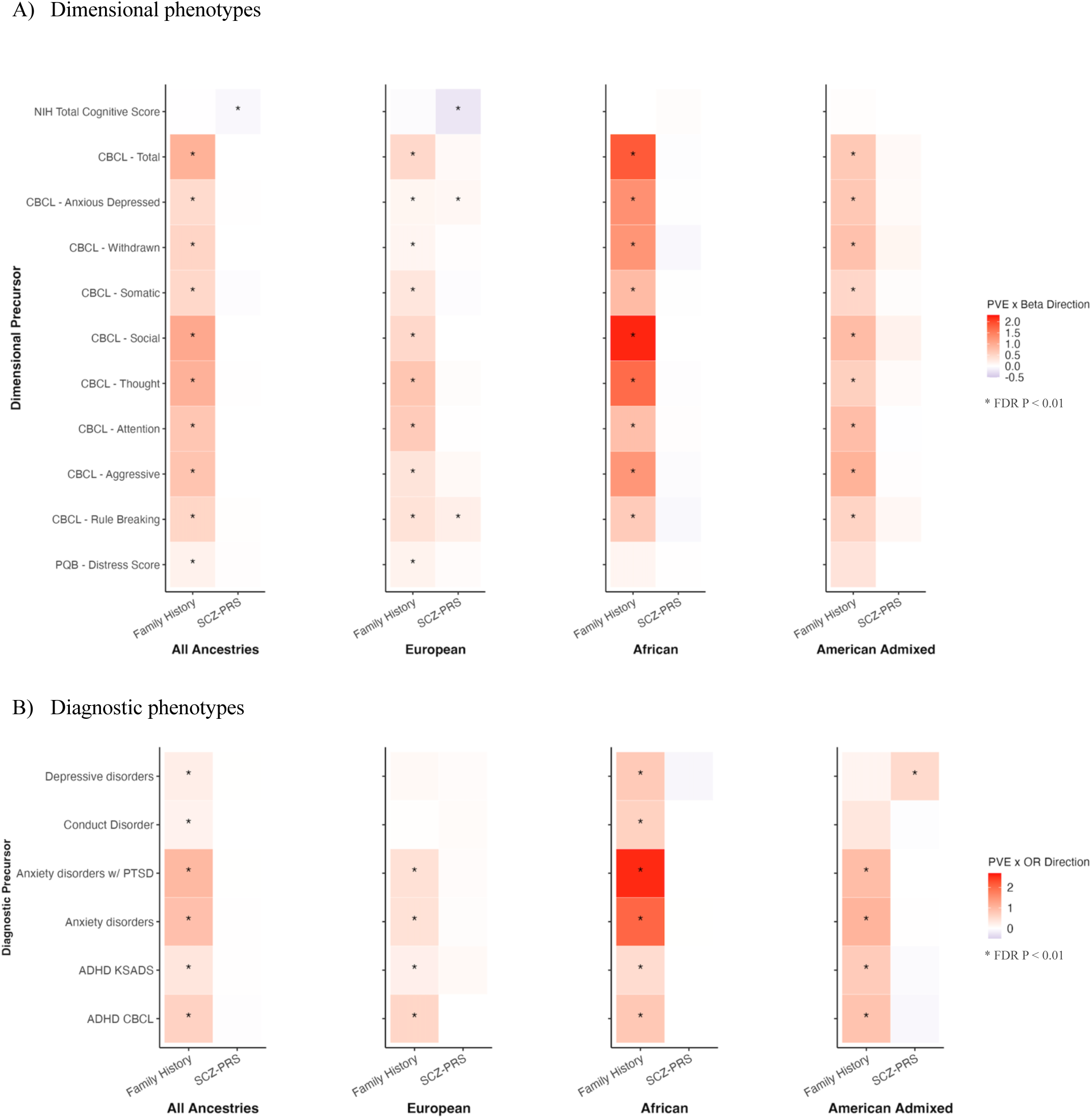
Heatmap of joint associations of schizophrenia polygenic risk scores (SCZ-PRS) and family history of psychosis (SCZ-FH) with (A) total cognitive score derived from the NIH-Toolbox, Child Behavior Checklist (CBCL) scores, and prodromal questionnaire (PQB) scores, and (B) lifetime history of psychiatric diagnoses derived from the Kiddie Schedule for Affective Disorders and Schizophrenia (KSADS). Heatmap shows associations of one genetic risk measure (i.e. SCZ-PRS or SCZ-FH) while adjusting for the other. FDR P refers to the false discovery rate–corrected p value.

**Table 3.**
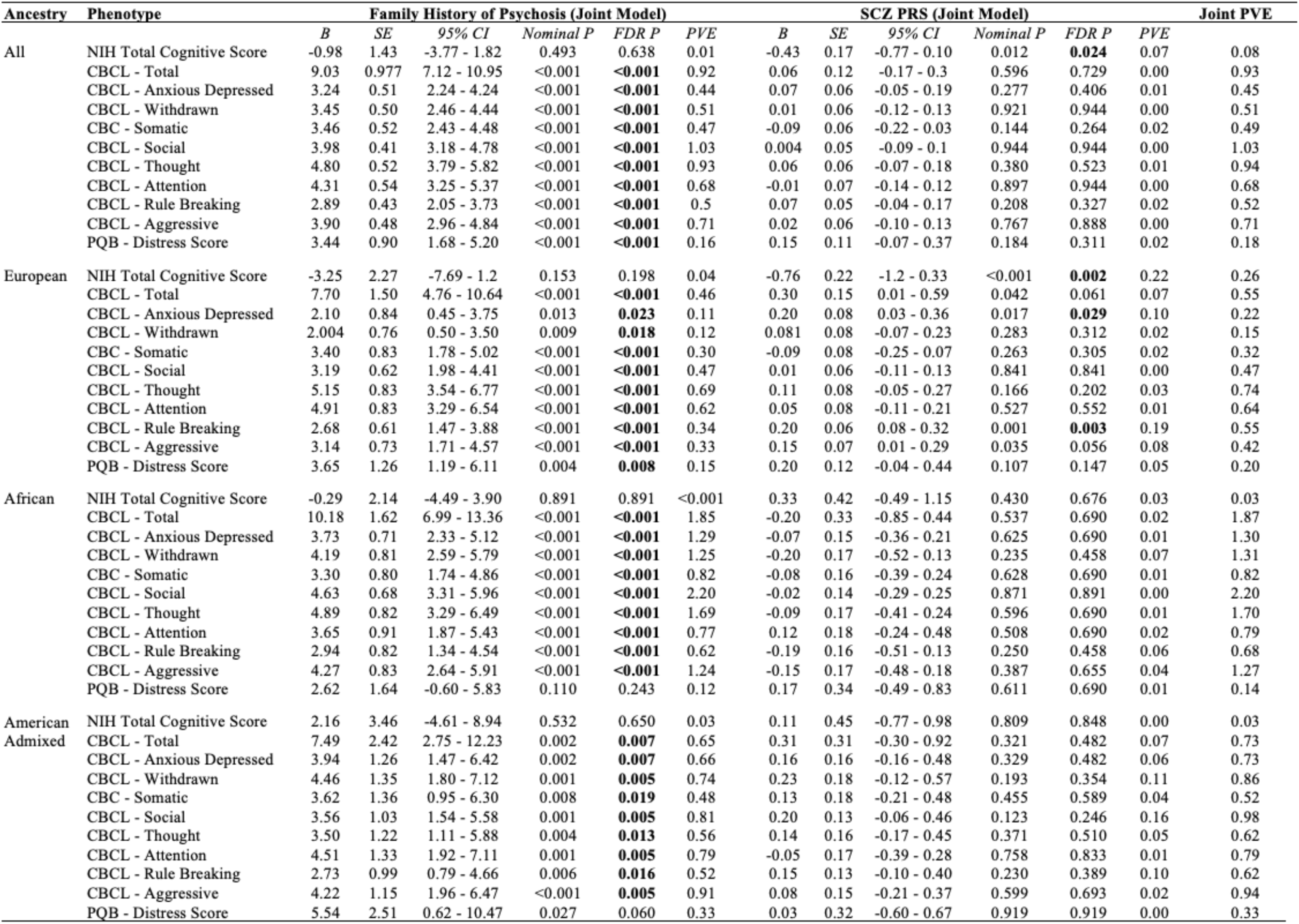
Joint model associations of schizophrenia polygenic risk scores (SCZ-PRS) and family history of psychosis (SCZ-FH) with dimensional, total cognitive score derived from the NIH-Toolbox, Child Behavior Checklist (CBCL) scores, and prodromal questionnaire (PQB) scores. FDR P refers to the false discovery rate–corrected p value; PVE refers to the percentage of variance explained.

SCZ-FH remained unassociated with the NIH-TB total cognition scores in the cross-ancestry analysis (*b*=-0.98, p=0.49, FDR p=0.64) or within individual ancestry groups, while higher SCZ-PRS was significantly associated with lower cognitive scores cross-ancestry (*b*= −0.43, p=0.01, FDR p=0.024) and for EUR-only (*b*=-0.76, p<0.001, FDR p=0.002) even after adjusting for SCZ-FH.

The joint model showed strong associations between SCZ-FH and CBCL total problems in the cross-ancestry analysis (*b*=9.03, p<0.001, FDR p<0.001) and within the three ancestry groups. Consistent with the independent model, SCZ-FH retained robust links with the 8 CBCL subscales in the cross-ancestry groups (*b* range=2.89-4.80, p<0.001, FDR p<0.001) and for EUR-only, AFR-only and AMR-only groups after adjusting for SCZ-PRS. SCZ-PRS was not associated with CBCL total problems (*b*=0.06, p=0.60, FDR p=0.73) or any of the 8 subscale scores (*b* range=-0.09-0.07, p=0.14-0.94, FDR p=0.26-0.94) in the cross-ancestry analysis. For EUR-only, associations of SCZ-PRS with CBCL anxious/depressed symptoms (*b*=0.20, p=0.02, FDR p=0.03) and rule breaking behavior (*b*=0.20, p=0.001, FDR p=0.003) scores remained significant, whereas the associations with total problems and aggressive behavior were no longer significant after accounting for SCZ-FH.

Similar to when modeled alone, SCZ-FH remained significantly associated with PQB Distress scores in the cross-ancestry analysis (b=3.44, p<0.001, FDR p<0.001) and for EUR-only youth (b=3.65, p=0.004, FDR p=0.01), after accounting for SCZ-PRS, while the association did not survive correction for AMR-only youth (b=5.54, p=0.03, FDR p=0.06) and was null in AFR-only youth (b=2.62, p=0.11, FDR p=0.24). Similar to when SCZ-PRS was modeled alone, SCZ-PRS was not associated with PQB Distress scores in the cross-ancestry group (*B*=0.15, p=0.18, FDR p=0.31) or within the three ancestry groups in models that simultaneously accounted for SCZ-FH.

### Joint associations between SCZ Risk Measures & Diagnostic Measures

Table 4 and Figure 1B summarize associations between the SCZ risk measures and clinical diagnoses in the joint models. Results were similar to the independent models. In the cross-ancestry analysis, greater SCZ-FH remained uniquely associated with higher likelihood of a depressive disorder (*OR*=2.94, p<0.001, FDR p<0.001), conduct disorder (*OR*=2.17, p<0.001, FDR p<0.001), anxiety disorder excluding (*OR*=4.74, p<0.001, FDR p<0.001), or including PTSD (*OR*=5.07, p<0.001, FDR p<0.001), and ADHD (*OR*=2.72, P<0.001, FDR p<0.001). Results were similar using CBCL-based current ADHD (*OR*=4.42, p<0.001, FDR p<0.001). As was the case in the independent model, associations of SCZ-FH with depressive and conduct disorder did not survive correction for multiple testing for EUR-only or AMR-only. SCZ-PRS was also not significantly associated with history of any clinical diagnoses in the cross-ancestry analysis or within specific ancestries after accounting for SCZ-FH, except for depressive disorders, for which the association with SCZ-PRS among AMR-only remained significant (*OR*=1.37, p=0.01, FDR p=0.02).

**Table 4.** Joint model associations of schizophrenia polygenic risk scores (SCZ-PRS) and family history of psychosis (SCZ-FH) with diagnostic phenotypes derived from the Kiddie Schedule for Affective Disorders and Schizophrenia (KSADS) or Child Behavior Checklist (CBCL). FDR P refers to the false discovery rate–corrected p value; PVE refers to the percentage of variance explained.

| Ancestry | Phenotype | Family History of Psychosis (Joint Model) |  |  |  |  |  | SCZ PRS (Joint Model) |  |  |  |  |  | Joint PVE |
| --- | --- | --- | --- | --- | --- | --- | --- | --- | --- | --- | --- | --- | --- | --- |
|  |  | OR | SE | 95% CI | Nominal P | FDR P | PVE | OR | SE | 95% CI | Nominal P | FDR P | PVE |  |
| All | KSADS - ADHD | 2.72 | 0.19 | 1.87 - 3.97 | <0.001 | **<0.001** | 0.35 | 1.03 | 0.03 | 0.97 - 1.08 | 0.331 | 0.497 | 0.01 | 0.36 |
|  | CBCL - ADHD | 4.42 | 0.23 | 2.79 - 7.01 | <0.001 | **<0.001** | 0.63 | 0.95 | 0.04 | 0.88 - 1.03 | 0.252 | 0.433 | 0.02 | 0.65 |
|  | KSADS - Anxiety Disorders | 4.74 | 0.20 | 3.20 - 7.04 | <0.001 | **<0.001** | 0.88 | 1.02 | 0.03 | 0.96 - 1.09 | 0.435 | 0.557 | 0.01 | 0.89 |
|  | KSADS - Anxiety disorders w/ PTSD | 5.07 | 0.20 | 3.44 - 7.50 | <0.001 | **<0.001** | 0.99 | 1.02 | 0.03 | 0.96 - 1.08 | 0.465 | 0.557 | 0.01 | 1.00 |
|  | KSADS - Depressive Disorders | 2.94 | 0.27 | 1.75 - 4.96 | <0.001 | **<0.001** | 0.24 | 1.03 | 0.05 | 0.94 - 1.13 | 0.510 | 0.557 | 0.01 | 0.25 |
|  | KSADS - Conduct Disorder | 2.17 | 0.21 | 1.43 - 3.30 | <0.001 | **<0.001** | 0.18 | 1.01 | 0.03 | 0.95 - 1.07 | 0.754 | 0.754 | 0.00 | 0.18 |
| European | KSADS - ADHD | 2.72 | 0.32 | 1.47 - 5.07 | 0.002 | **0.005** | 0.21 | 1.08 | 0.03 | 1.01 - 1.15 | 0.034 | 0.083 | 0.09 | 0.32 |
|  | CBCL - ADHD | 6.67 | 0.40 | 3.06 - 14.55 | <0.001 | **<0.001** | 0.57 | 1.01 | 0.06 | 0.90 - 1.13 | 0.875 | 0.875 | 0.00 | 0.57 |
|  | KSADS - Anxiety Disorders | 3.95 | 0.32 | 2.11 - 7.40 | <0.001 | **<0.001** | 0.41 | 1.05 | 0.04 | 0.98 - 1.13 | 0.168 | 0.202 | 0.04 | 0.47 |
|  | KSADS - Anxiety disorders w/ PTSD | 3.97 | 0.32 | 2.12 - 7.41 | <0.001 | **<0.001** | 0.42 | 1.06 | 0.04 | 0.99 - 1.14 | 0.116 | 0.175 | 0.05 | 0.49 |
|  | KSADS - Depressive Disorders | 2.78 | 0.54 | 0.96 - 8.08 | 0.060 | 0.119 | 0.09 | 1.11 | 0.07 | 0.97 - 1.26 | 0.134 | 0.179 | 0.05 | 0.15 |
|  | KSADS - Conduct Disorder | 1.37 | 0.36 | 0.67 - 2.79 | 0.384 | 0.419 | 0.02 | 1.06 | 0.04 | 0.99 - 1.15 | 0.094 | 0.161 | 0.06 | 0.08 |
| African | KSADS - ADHD | 2.34 | 0.29 | 1.33 - 4.12 | 0.003 | **0.006** | 0.50 | 1.008 | 0.07 | 0.89 - 1.15 | 0.900 | 0.973 | 0.00 | 0.50 |
|  | CBCL - ADHD | 3.07 | 0.33 | 1.60 - 5.90 | <0.001 | **0.003** | 0.78 | 0.98 | 0.09 | 0.82 - 1.16 | 0.778 | 0.973 | 0.00 | 0.78 |
|  | KSADS - Anxiety Disorders | 5.11 | 0.31 | 2.80 - 9.31 | <0.001 | **<0.001** | 2.04 | 1.024 | 0.08 | 0.87 - 1.20 | 0.770 | 0.973 | 0.01 | 2.05 |
|  | KSADS - Anxiety disorders w/ PTSD | 6.14 | 0.30 | 3.40 - 11.08 | <0.001 | **<0.001** | 2.57 | 0.997 | 0.08 | 0.85 - 1.16 | 0.973 | 0.973 | 0.00 | 2.57 |
|  | KSADS - Depressive Disorders | 3.21 | 0.35 | 1.61 - 6.41 | <0.001 | **0.003** | 0.74 | 0.887 | 0.1 | 0.73 - 1.08 | 0.224 | 0.384 | 0.09 | 0.83 |
|  | KSADS - Conduct Disorder | 2.66 | 0.30 | 1.47 - 4.81 | 0.001 | **0.003** | 0.64 | 1.016 | 0.08 | 0.88 - 1.18 | 0.830 | 0.973 | 0.00 | 0.64 |
| American<br>Admixed | KSADS - ADHD | 4.18 | 0.48 | 1.62 - 10.78 | 0.003 | **0.009** | 0.73 | 0.95 | 0.07 | 0.82 - 1.10 | 0.462 | 0.616 | 0.05 | 0.76 |
|  | CBCL - ADHD | 6.05 | 0.60 | 1.87 - 19.51 | 0.003 | **0.009** | 0.84 | 0.91 | 0.11 | 0.73 - 1.12 | 0.359 | 0.539 | 0.08 | 0.90 |
|  | KSADS - Anxiety Disorders | 5.77 | 0.52 | 2.07 - 16.08 | <0.001 | **0.009** | 1.05 | 1.04 | 0.09 | 0.87 - 1.24 | 0.682 | 0.744 | 0.01 | 1.07 |
|  | KSADS - Anxiety disorders w/ PTSD | 5.24 | 0.52 | 1.88 - 14.59 | 0.002 | **0.009** | 0.93 | 1.01 | 0.09 | 0.85 - 1.20 | 0.905 | 0.905 | 0.00 | 0.93 |
|  | KSADS - Depressive Disorders | 2.70 | 0.72 | 0.66 - 11.03 | 0.167 | 0.286 | 0.15 | 1.37 | 0.12 | 1.08 - 1.73 | 0.009 | **0.021** | 0.52 | 0.68 |
|  | KSADS - Conduct Disorder | 3.17 | 0.57 | 1.03 - 9.77 | 0.044 | 0.089 | 0.34 | 0.95 | 0.09 | 0.79 - 1.15 | 0.601 | 0.722 | 0.02 | 0.36 |

### Sensitivity Analysis Incorporating Income-to-Needs

When controlling for income-to-needs, most associations remained consistent with the main analyses, although some SCZ-FH-related associations no longer reached significance (Tables S2–S5). In both independent and joint models, SCZ-FH was no longer associated with CBCL attention and rule-breaking symptoms in the AFR-only model, or with somatic and rule-breaking symptoms in AMR-only youth. In the joint model, SCZ-FH was no longer associated with CBCL total and withdrawn symptom severity in AMR-only youth. For diagnostic phenotypes, adjusting for income-to-needs in the independent models attenuated associations between SCZ-FH and conduct disorders in AFR-only and AMR-only youth, as well as ADHD diagnoses in AFR-only youth. In contrast, associations with lifetime clinical diagnoses remained largely unchanged in the joint models. These findings suggest that poverty-related factors may partially account for SCZ-FH associations with psychopathology, indicating that SCZ-FH may capture risk beyond genetic predispositions including environmental influences. The stability of SCZ-PRS associations across models may suggest a more direct genetic contribution underscoring partially distinct pathways of risk for SCZ-FH and SCZ-PRS.

## Discussion

This study demonstrates partially distinct and independent associations of SCZ-FH and SCZ-PRS with cognitive, behavioral, and emotional functioning in childhood in the ABCD study. SCZ-FH showed broad cross-ancestry associations with dimensional and diagnostic markers of psychopathology without association to cognitive functioning. In contrast, SCZ-PRS showed more specific associations to lower cognitive functioning across ancestries, and to total and anxious-depressed symptoms, as well as rule-breaking and aggressive behaviors within the EUR-only group. Notably, when both SCZ-FH and SCZ-PRS were included in joint models, SCZ-FH retained robust associations across dimensional and diagnostic measures, while SCZ-PRS effects on cognition and specific behavioral and emotional symptoms persisted, underscoring their relatively independent associations with functioning.

Across ancestries, greater SCZ-FH was linked with greater emotional and behavioral problems, more distressing childhood psychotic-like experiences, and higher likelihood of clinical diagnoses, including depressive, conduct, and anxiety disorders, and ADHD. These findings align with previous ABCD-specific findings which showed that a binary measure of SCZ-FH was associated with internalizing and externalizing problems, higher psychotic-like symptom and KSADS reports of ADHD and PTSD among youth of European ancestry (Loughnan et al., 2022). Interestingly, this prior study identified associations between SCZ-FH and KSADS-based depressive and conduct disorders in European ancestry youth after controlling for SES, which we did not observe in our sensitivity analysis. Conversely, our study found associations between SCZ-FH and anxiety disorders, a relationship not reported by Loughnan et al. (2022). These incongruent findings likely reflect differences in the operationalization of SCZ-FH and socioeconomic status (SES). Both studies used the same question to assess SCZ-FH, but we employed a weighted measure that accounted for number of affected relatives and degree of relatedness. Similarly, we defined SES using income-to-needs ratio whereas this prior study used parental education and household income. Nevertheless, current results highlight robust associations between greater SCZ-FH and emotional, behavioral, and clinical problems in childhood, and suggest that a weighted SCZ-FH measure may help uncover associations with some diagnoses not observed previously.

Associations between SCZ-PRS and childhood psychopathology were only detected within specific ancestries. In the EUR-only models, higher SCZ-PRS was linked to increased severity of CBCL total problems, anxious/depressed, rule breaking and aggressive behavior. In AMR-only models, SCZ-PRS was associated with depressive diagnoses. Although SCZ-PRS was associated with some CBCL variables in EUR-only, the links were weaker compared to associations between CBCL scores and SCZ-FH. Nevertheless, we observed more significant associations with emotional and behavioral problems in ABCD study youth at baseline than reported in some recent studies. For example, Wainberg et al. (2022) did not find significant associations between SCZ-PRS and the eight CBCL subscales in unrelated European ancestry youth, while Loughnan et al. (2022) noted a significant relationship only with CBCL Rule-Breaking. This may be due to our use of a GRM to account for relatedness, enabling a larger sample than studies which excluded related individuals. Furthermore, differences in PRS generation methods, with our study employing PRS-CSx may have facilitated our ability to detect these associations. Overall, SCZ-PRS showed weak but detectable associations with childhood psychopathology, suggesting modest effects of molecularly defined genetic risk for SCZ on psychopathology in childhood.

Cognitive functioning in childhood was not associated with SCZ-FH but was linked to SCZ-PRS in the cross-ancestry analysis and in the EUR-only group. This finding that higher SCZ-PRS predicts poorer cognitive performance aligns with prior research (Loughnan et al., 2022; Mollon & Reichenberg, 2018), but the lack of association with SCZ-FH contrasts prior findings that offspring of parents with psychosis exhibit cognitive deficits (MacKenzie et al., 2020; Seidman, 2010). Weaker associations of SCZ-FH with cognition in ABCD may reflect the normative characteristics of the sample, reliance on a single-question SCZ-FH measure, and/or the use of the NIH-TB over traditional neurocognitive assessments like the Wechsler Intelligence Scale for Children (WISC). Nevertheless, our results suggest that poorer cognitive function may more directly reflect consequences of genetic risk for SCZ than broader aspects of SCZ risk that are captured by SCZ-FH.

The effect sizes and significance levels for SCZ risk predictors in the joint models closely mirrored those in the independent models, suggesting that SCZ-FH and SCZ-PRS capture partially distinct aspects of risk for SCZ. This aligns with growing evidence that SCZ-FH and PRS offer independent yet complementary insights into disease susceptibility for SCZ and other conditions (Loughnan et al., 2022; Lu et al., 2018; Mars et al., 2022). SCZ-FH may capture environmental aspects of risk, such as intergenerational exposure to stress, or increased symptom recognition in families with a history of SCZ (Bowers & Yehuda, 2016; Oliver-Parra, Dalmau-Bueno, Ruiz-Muñoz, & García-Altés, 2020). In line with this, some associations between SCZ-FH and psychopathology symptoms were attenuated after accounting for income-to-needs ratio, whereas the narrower associations between SCZ-PRS and cognitive functioning, as well as subset of psychopathology symptoms in European ancestry youth, were unchanged. Together, these findings highlight the potential clinical value of combining SCZ-FH and SCZ-PRS for early risk assessment.

Although we identified associations between the SCZ-PRS and aspects of cognitive, emotional, and behavioral functioning in European ancestry youth, the strength of SCZ-PRS associations in non-European ancestry groups were weaker. These findings underscore the importance of improving PRS accuracy across groups through increased GWAS representation, continued PRS methods development, and establishment of clear standards for PRS analyses. For example, our use of within-ancestry z-scoring aligns with a common scaling method for creating PRS clinical cutoffs or odds ratios across groups but this is not a definitive standard in the field (Khan et al., 2022). Conceptual advancements for how to control for ancestry differences, such as using covariates reflecting genetic distance to original GWAS samples, are promising but methods for implementation are still developing (Ding et al., 2023). In summary, while PRS methodology and inclusion efforts have advanced, ongoing research is needed to establish robust and inclusive standards for cross-ancestry PRS analyses to enhance our understanding of genetic-mediated precursors for disorders.

### Limitations

One limitation is the reliance on parent-report CBCL and KSADS-COMP measures, which, while generally showing stronger concurrent validity with gold-standard clinician interviews for diagnoses and symptom severity at this age (Townsend et al., 2020; Warnick, Bracken, & Kasl, 2008), may introduce bias due to subjective perceptions (Robinson et al., 2019). Second, the psychometric properties of the overall cognitive functioning measure derived from the NIH-TB are less robust than some traditional measures of cognitive functioning, such as the WISC (Taylor et al., 2022). However, the NIH-TB has notable advantages, including ease of administration which improves scalability and predictive validity, making it a reasonably effective measure of cognition (Distefano et al., 2023). Additionally, although the ABCD study aimed at reflecting general U.S. demographics, a general population sample may also be lower powered to detect associations between SCZ risk and aspects of functioning because of low base rate of SCZ compared to samples enriched for SCZ risk.

## Conclusion

In summary, the current study identified strong associations between SCZ-FH and psychopathology but not cognitive functioning in the ABCD Study, while SCZ-PRS showed more variable associations, with stronger links to cognition across ancestries and specific emotional and behavioral symptoms in European ancestry youth. These associations remained largely unchanged when SCZ-FH and SCZ-PRS were jointly modeled, underscoring the complementary information to be gained from considering both risk measures. Nevertheless, associations between SCZ-PRS and cognitive and emotional functioning in non-European ancestry youth were substantially weaker compared to European ancestry youth. Clarifying whether this reflects lower SCZ-PRS accuracy or greater influence of other risk factors on childhood functioning in diverse populations is crucial. Future research should prioritize inclusive methodologies to improve understanding of SCZ risk and childhood functioning across diverse youth, and longitudinal studies to explore the temporal progression of these relationships.

## Data Availability

All data produced in the present study are available upon reasonable request to the authors.

## Acknowledgments

Data used in the preparation of this manuscript were obtained from the Adolescent Brain Cognitive Development (ABCD) Study (https://abcdstudy.org), held in the National Institute of Mental Health (NIMH) Data Archive (NDA). NDA is a collaborative informatics system created by the National Institutes of Health to provide a national resource to support and accelerate research in mental health. Dataset identifier(s): https://doi.org/10.15154/1523041. This study was also funded by the University of Washington Population Health Initiative Tier 1 Pilot Research Grant (MH, AEF, KTF, JKF).

