## Supplemental Methods and Figures for "Independent Versus Joint Effects of Polygenic or Family-Based Schizophrenia Risk in Diverse Ancestry Youth in the ABCD Study"

### Supplementary Information

#### Supplementary Methods

Measures

*SCZ-related Signs and Symptoms*
I) Dimensional Assessments
A. NIH Cognitive Toolbox

Children completed tasks from the NIH Cognitive Toolbox (NIH-TB) which quantifies cognitive functioning across various domains (Gershon et al., 2013; Weintraub et al., 2013). The tasks include Flanker (inhibitory control), List Sorting (working memory), Picture Sequence (episodic memory), Oral Reading Recognition (language), Picture Vocabulary (language), and Pattern Comparison (processing speed) subtests. The Flanker task tests participants' ability to inhibit attention from irrelevant stimuli. List Sorting requires the participant to recall and sequence different stimuli while Picture Sequence is a test of episodic memory and involves the reproduction of pictures. Both Oral Reading Recognition and Picture Vocabulary assess language skills with the former requiring participants to read letters and words aloud, while the latter involves selecting the most fitting picture for a word presented audibly. Lastly, Pattern Comparison measures processing speed by requiring participants to quickly determine whether two stimuli are the same. A total composite, age-corrected cognitive score, summarizing functioning across these tasks was used for analysis, after winsorizing scores to account for outliers beyond three standard deviations.

B. Child Behavior Checklist

Primary caregivers completed the CBCL which is a well-validated and widely used assessment involving 113 questions to measure emotional and behavioral problems in children over the past 6 months (Achenbach, 1991). The total composite score, along with scores for the eight primary subscales: anxious/depressed, withdrawn/depressed, somatic complaints, social problems, thought problems, attention problems, rule-breaking behavior, and aggressive behavior severity were used for analysis.

C. Prodromal Questionnaire – Brief Child Version

Children completed the Prodromal Questionnaire – Brief Child Version (PQ-BC) which is a 21-item scale that assesses the occurrences of psychotic-like experiences (e.g. perceptual abnormalities), and level of distress for any endorsed experience. We generated a summary PQ-BC Distress score by the adding the total number of endorsements weighted by distress (for each item, 0 = did not experience PLE, 1 = experienced PLE with no distress, 2-6 = distressing PLE [+1 to the distress score for the item]) (Chang et al., 2024). The PQ-BC Distress score was used as the measure of psychotic-like experiences (PLEs) for analysis.

II) Diagnostic Assessments
A. Kiddie Schedule for Affective Disorders and Schizophrenia

Caregivers and youth completed a semi-structured, self-administered, computerized version of the KSADS-5 which assesses mental health conditions experienced by the child, including depression, bipolar disorder, anxiety disorders, psychosis, conduct disorder and ADHD (KSADS-COMP; Kaufman et al., 1997). Lifetime history of depressive disorders, conduct disorder, anxiety disorders (with and without post-traumatic stress disorder) and attention-deficit/hyperactivity disorder (ADHD) diagnoses were derived from the caregiver self-administered KSADS-COMP, as they have been shown to have greater concordance with diagnoses derived from gold-standard clinician interviews integrating parent and youth report, compared to diagnoses derived from the youth self-administered KSADS-COMP (Townsend et al., 2020). For the diagnosis of depressive disorders, lifetime disruptive mood dysregulation disorder, major depressive disorder, or persistent depressive disorder, were included. For the diagnosis of anxiety disorders, agoraphobia, generalized anxiety disorder, panic, separation anxiety, and social anxiety disorder, were included. An additional metric for lifetime history of anxiety disorders including post-traumatic stress disorder (PTSD), was also analyzed. To facilitate comparison with dimensional psychopathology measures derived from the CBCL, analyses of KSADS-derived clinical diagnoses utilized lifetime history of any depressive disorder, any anxiety disorder (including and excluding PTSD), conduct disorder, and ADHD. Potential issues with KSADS ADHD diagnoses in ABCD have been noted previously due to a modification of the diagnostic criterion to require impairment in two or more settings, which yielded an unexpectedly low endorsement rate (Barch et al., 2021). To address this, we re-coded the variable to include cases with impairment in just one setting, an approach that aligns with the criterion originally used in Data Release 3.

B. CBCL ADHD variable

To facilitate comparison with alternate methods for deriving ADHD diagnoses, we generated an additional ADHD variable using CBCL cut-offs. A T-score of ≥65 on the CBCL Attention Problems scale was established as the threshold for clinical-level attention problems. A key distinction between the diagnostic measures is that the KSADS variables reflect the lifetime prevalence of ADHD, while the CBCL-derived ADHD measure captures symptoms occurring only within the past six months.

#### Statistical Analysis

##### *Ancestry Principal Components, Ancestry Grouping, and Genetic Relatedness*

To create ancestry groups of subjects, we conducted principal components analysis (PCA) using high-quality SNPs from the ABCD dataset merged with HapMap3 data as the reference (The International HapMap 3 Consortium, 2010). HapMap3 includes genotype data from 11 global populations, providing a robust reference for ancestry analysis. To ensure inclusion of only high-quality SNPs, we excluded variants that violated Hardy-Weinberg Equilibrium at a p-threshold of 0.001 during the merging process (Chang et al., 2024). Ancestry representative PCs were generated following an iterative procedure optimized for samples from diverse ancestral groups with familial relatedness (Conomos, Laurie, et al., 2016). This involved using PC-AiR for PC calculation and PC-Relate for kinship coefficient estimation, implemented in the GENESIS R package (Conomos, Miller, & Thornton, 2015; Conomos, Reiner, Weir, & Thornton, 2016).

We utilized these ancestry PCs as inputs for a Random Forest classifier to categorize participants into ancestry groups based on genetic similarity, using HapMap3 super-population labels (Alexander & Lange, 2011). The top eight PCs, which captured the majority of ancestry variance, were used to train the classifier. A probability threshold of 0.7 was applied for ancestry group assignment, leading to a slight reduction in the sample size from 10,979 to 9,365 participants but better grouping accuracy. Figure S1 provides a visualization of the variance explained by the top 8 PCs.

*Association Testing*

To optimize computational efficiency, the GRMs were made sparse by setting a fourth-degree relatedness threshold; values in the GRM below this threshold were set to 0. We utilized the fitNullModel function within the GENESIS package to perform linear and logistic regressions, as it accommodates matrix covariates (e.g. the GRM) as random effects. P-values for all dimensional measures were corrected for multiple testing using false discovery rate (FDR) correction within each ancestry group, across both risk SCZ measures. P-values for all diagnostic outcomes were similarly FDR-corrected for multiple testing within each ancestry group, across both risk SCZ measures. We also conducted a sensitivity analysis incorporating the income-to-needs ratio as an additional covariate. This ratio was calculated by dividing the median income value within each income band by the federal poverty threshold, adjusted for household size (Hair et al., 2015).

### Supplementary Tables & Figures

Figure S1. Scatterplot of the top 8 ancestry PCs, including the ABCD and HapMap3 reference samples. CEU: Utah residents with Northern and Western European ancestry; CHB: Han Chinese in Beijing, China; YRI: Yoruba in Ibadan, Nigeria; TSI: Toscans in Italy; JPT: Japanese in Tokyo, Japan; CHD: Chinese in Metropolitan Denver, Colorado; MEX: Mexican ancestry in Los Angeles, California; GIH: Gujarati Indians in Houston, Texas; ASW: Gujarati Indians in Houston, Texas; LWK: Luhya in Webuye, Kenya; MKK: Maasai in Kinyawa, Kenya

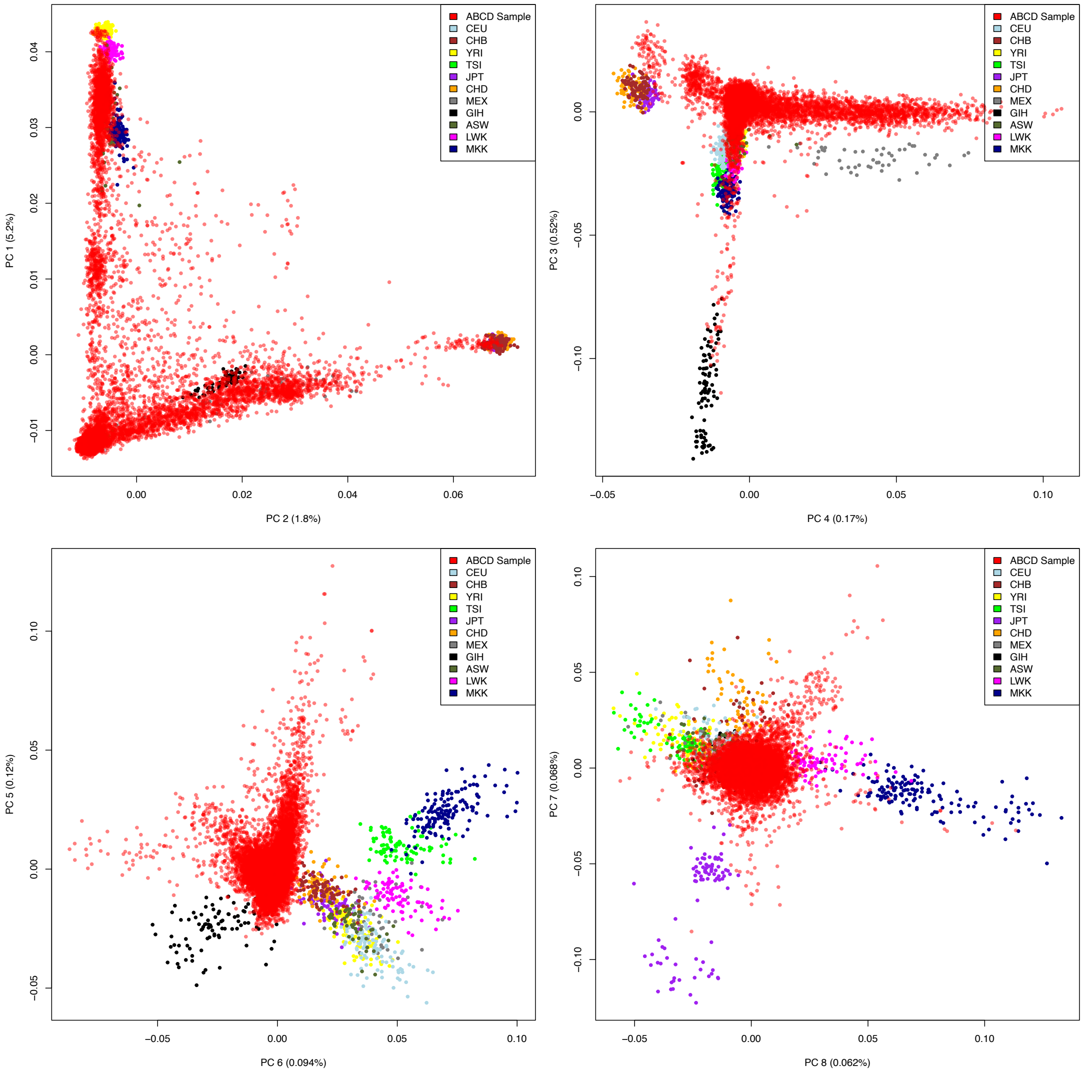

Figure S2. Scatterplot of associations between schizophrenia polygenic risk scores (SCZ-PRS) and family history of psychosis (SCZ-FH) for each of the three ancestries. EUR-only refers to European ancestry youth; AFR-only refers to African ancestry youth; AMR refers to American Admixed ancestry youth

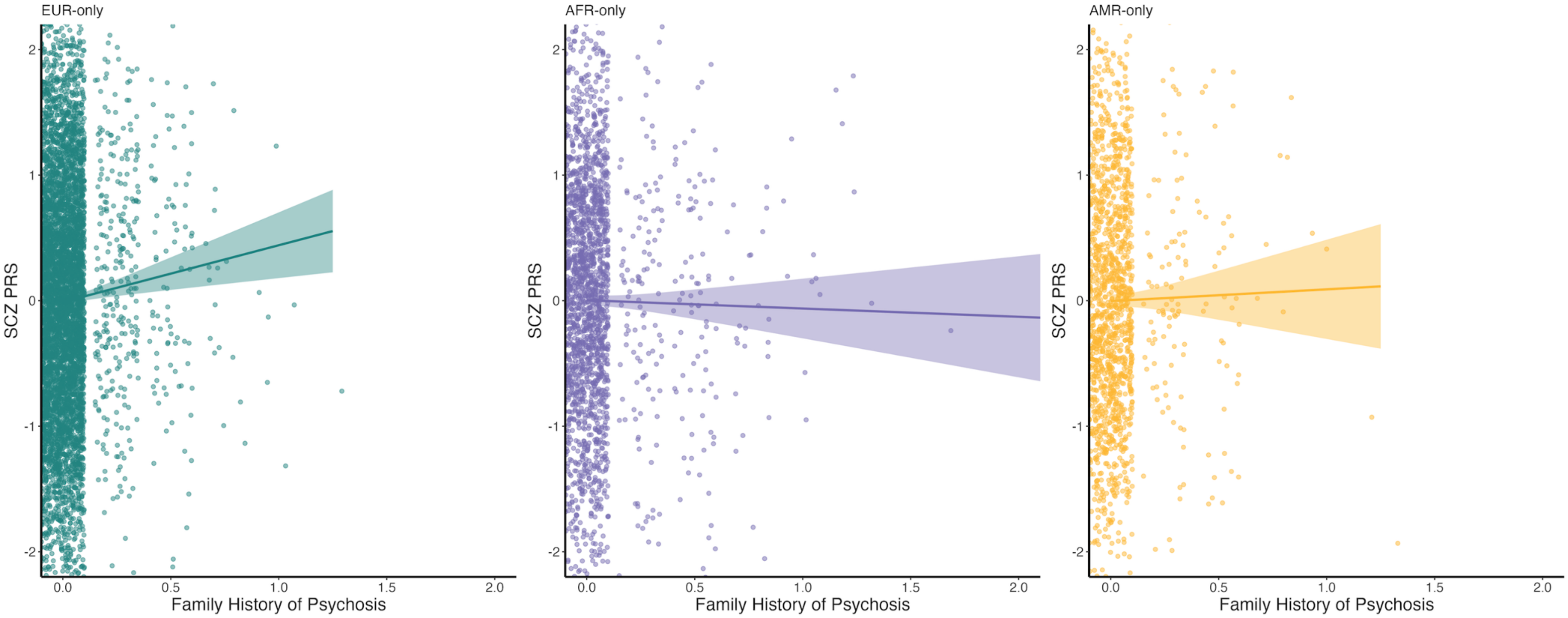

Figure S3. Heatmap of independent associations of schizophrenia polygenic risk scores (SCZ-PRS) and family history of psychosis (SCZ-FH) with total cognitive score derived from the NIH-Toolbox, Child Behavior Checklist (CBCL) scores, and prodromal questionnaire (PQB) scores. FDR P refers to the false discovery rate–corrected p value.

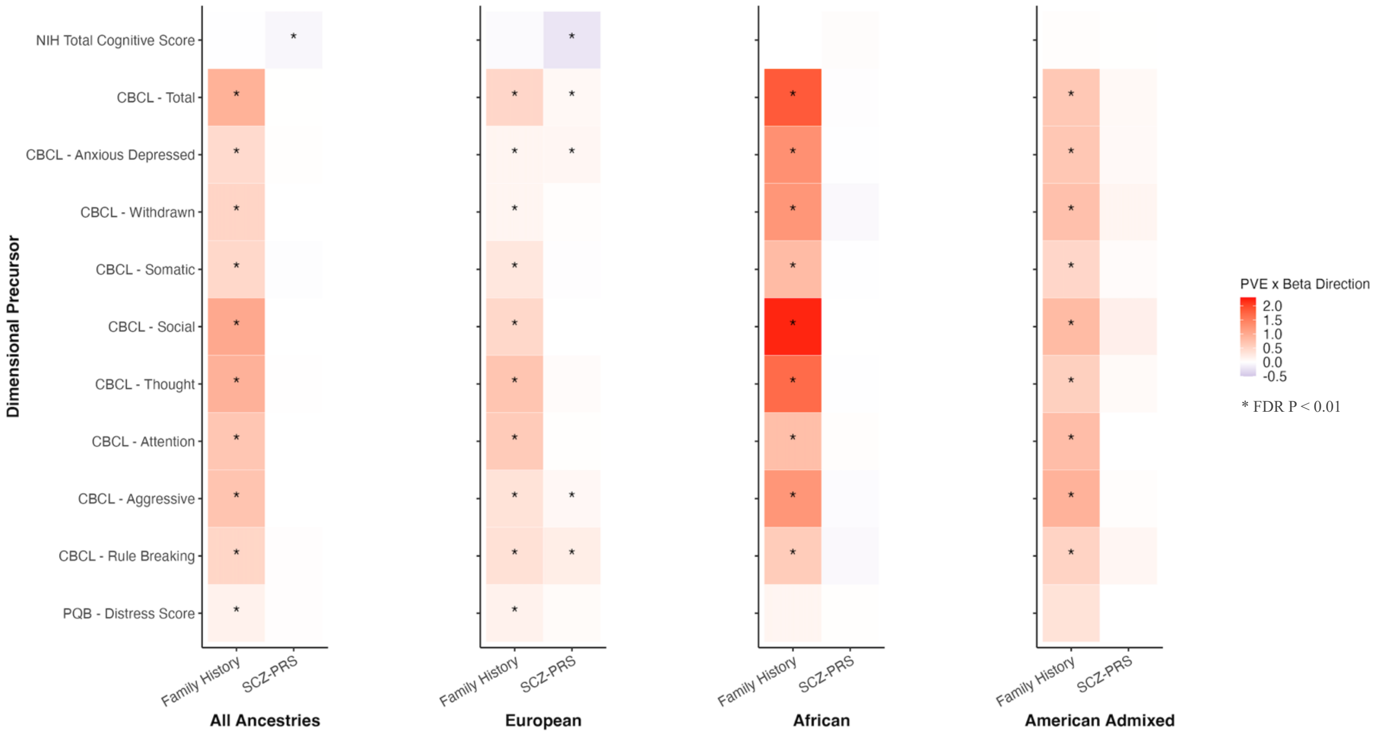


Figure S4. Heatmap of independent associations of schizophrenia polygenic risk scores (SCZ-PRS) and family history of psychosis (SCZ-FH) with lifetime history of psychiatric diagnoses derived from the Kiddie Schedule for Affective Disorders and Schizophrenia (KSADS). FDR P refers to the false discovery rate–corrected p value.

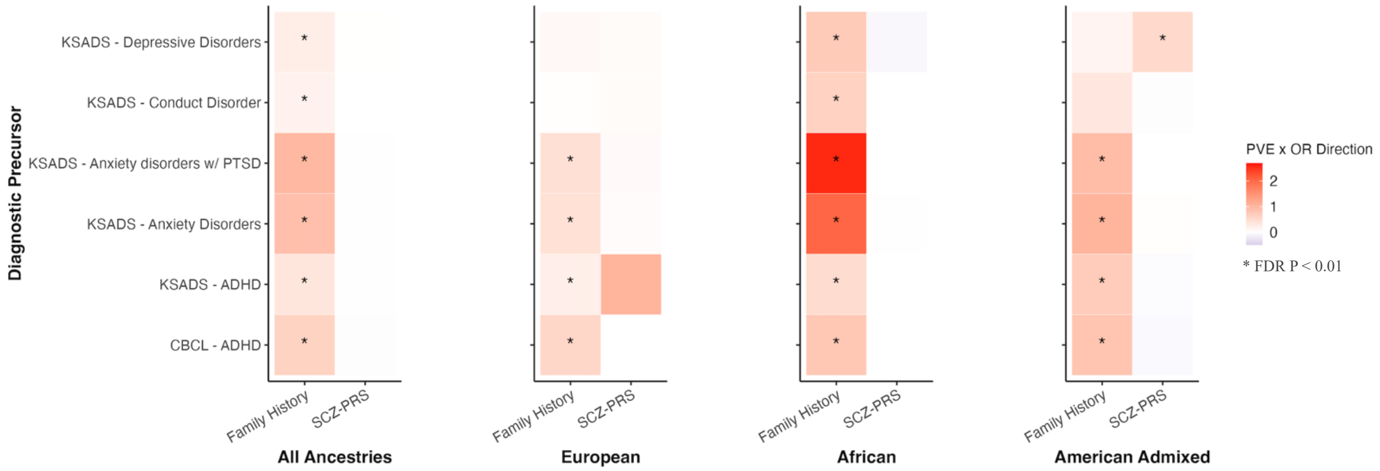

Table S1. The number of participants for whom data is available per phenotype measure for each ancestry group

| **Phenotype Measure** | **European** | **African** | **Admixed American** | **Across Ancestry (Total)** |
|---|---|---|---|---|
| NIH-TB Cognition | 5,452 | 2,020 | 1,425 | 8,897 |
| CBCL | 5,633 | 2,093 | 1,474 | 9,200 |
| PQB | 5,636 | 2,093 | 1,477 | 9,206 |
| KSADS-COMP | 5,621 | 2,087 | 1,471 | 9,179 |

Table S2. Independent associations of schizophrenia polygenic risk scores (SCZ-PRS) and family history of psychosis (SCZ-FH) with dimensional, total cognitive score derived from the NIH-Toolbox, Child Behavior Checklist (CBCL) scores, and prodromal questionnaire (PQB) scores, including income-to-needs as a covariate. FDR P refers to the false discovery rate–corrected p value; PVE refers to the percentage of variance explained.

| **Ancestry** | **Genetic Risk Measure** | **Phenotype** | **B** | **SE** | **95% CI** | **Nominal P** | **FDR P** | **PVE (%)** |
|---|---|---|---|---|---|---|---|---|
| All | Family History | NIH Total Cognitive Score | 0.83 | 1.49 | -2.08 - 3.75 | 0.5758 | 0.6333 | 0.00 |
| | | CBCL - Total | 7.19 | 1.02 | 5.20 - 9.18 | <0.001 | **<0.001** | 0.60 |
| | | CBCL - Anxious Depressed | 2.78 | 0.54 | 1.72 - 3.84 | <0.001 | **<0.001** | 0.32 |
| | | CBCL - Withdrawn | 2.10 | 0.52 | 1.08 - 3.12 | <0.001 | **<0.001** | 0.20 |
| | | CBC - Somatic | 2.81 | 0.55 | 1.72 - 3.89 | <0.001 | **<0.001** | 0.31 |
| | | CBCL - Social | 3.58 | 0.42 | 2.75 - 4.41 | <0.001 | **<0.001** | 0.85 |
| | | CBCL - Thought | 4.33 | 0.55 | 3.25 - 5.40 | <0.001 | **<0.001** | 0.75 |
| | | CBCL - Attention | 3.30 | 0.57 | 2.18 - 4.42 | <0.001 | **<0.001** | 0.40 |
| | | CBCL - Rule Breaking | 1.90 | 0.44 | 1.04 - 2.77 | <0.001 | **<0.001** | 0.22 |
| | | CBCL - Aggressive | 3.18 | 0.51 | 2.19 - 4.18 | <0.001 | **<0.001** | 0.47 |
| | | PQB - Distress Score | 3.25 | 0.94 | 1.40 - 5.09 | <0.001 | **0.001** | 0.14 |
| | SCZ PRS | NIH Total Cognitive Score | -0.48 | 0.18 | -0.82 - -0.13 | 0.007 | **0.014** | 0.09 |
| | | CBCL - Total | 0.17 | 0.12 | -0.08 - 0.41 | 0.178 | 0.245 | 0.02 |
| | | CBCL - Anxious Depressed | 0.10 | 0.07 | -0.03 - 0.23 | 0.129 | 0.189 | 0.03 |
| | | CBCL - Withdrawn | 0.03 | 0.06 | -0.09 - 0.16 | 0.616 | 0.646 | 0.00 |
| | | CBC - Somatic | -0.09 | 0.07 | -0.22 - 0.05 | 0.198 | 0.256 | 0.02 |
| | | CBCL - Social | 0.04 | 0.05 | -0.06 - 0.14 | 0.429 | 0.525 | 0.01 |
| | | CBCL - Thought | 0.10 | 0.07 | -0.03 - 0.23 | 0.125 | 0.189 | 0.03 |
| | | CBCL - Attention | -0.01 | 0.07 | -0.14 - 0.13 | 0.932 | 0.932 | 0.00 |
| | | CBCL - Rule Breaking | 0.12 | 0.05 | 0.01 - 0.22 | 0.029 | 0.054 | 0.06 |
| | | CBCL - Aggressive | 0.04 | 0.06 | -0.08 - 0.16 | 0.479 | 0.555 | 0.01 |
| | | PQB - Distress Score | 0.20 | 0.12 | -0.02 - 0.43 | 0.078 | 0.132 | 0.04 |
| European | Family History | NIH Total Cognitive Score | -0.75 | 2.31 | -5.27 - 3.77 | 0.744 | 0.779 | 0.00 |
| | | CBCL - Total | 6.38 | 1.53 | 3.38 - 9.37 | <0.001 | **<0.001** | 0.32 |
| | | CBCL - Anxious Depressed | 1.96 | 0.87 | 0.26 - 3.66 | 0.024 | **0.042** | 0.10 |
| | | CBCL - Withdrawn | 1.08 | 0.78 | -0.44 - 2.60 | 0.163 | 0.211 | 0.04 |
| | | CBC - Somatic | 2.64 | 0.85 | 0.97 - 4.31 | 0.002 | **0.005** | 0.18 |
| | | CBCL - Social | 2.71 | 0.63 | 1.48 - 3.95 | <0.001 | **<0.001** | 0.34 |
| | | CBCL - Thought | 4.89 | 0.85 | 3.23 - 6.55 | <0.001 | **<0.001** | 0.62 |
| | | CBCL - Attention | 4.32 | 0.85 | 2.64 - 5.99 | <0.001 | **<0.001** | 0.48 |
| | | CBCL - Rule Breaking | 2.26 | 0.62 | 1.04 - 3.48 | <0.001 | **<0.001** | 0.25 |
| | | CBCL - Aggressive | 2.88 | 0.74 | 1.42 - 4.33 | <0.001 | **<0.001** | 0.28 |
| | | PQB - Distress Score | 3.51 | 1.30 | 0.95 - 6.06 | 0.007 | **0.016** | 0.14 |
| | SCZ PRS | NIH Total Cognitive Score | -0.68 | 0.22 | -1.12 - -0.24 | 0.002 | **0.006** | 0.18 |
| | | CBCL - Total | 0.32 | 0.15 | 0.03 - 0.61 | 0.031 | **0.049** | 0.09 |
| | | CBCL - Anxious Depressed | 0.20 | 0.08 | 0.04 - 0.37 | 0.017 | **0.034** | 0.11 |
| | | CBCL - Withdrawn | 0.10 | 0.08 | -0.05 - 0.24 | 0.204 | 0.250 | 0.03 |
| | | CBC - Somatic | -0.09 | 0.08 | -0.25 - 0.08 | 0.304 | 0.352 | 0.02 |
| | | CBCL - Social | 0.01 | 0.06 | -0.11 - 0.13 | 0.839 | 0.839 | 0.00 |
| | | CBCL - Thought | 0.14 | 0.08 | -0.02 - 0.30 | 0.085 | 0.116 | 0.06 |
| | | CBCL - Attention | 0.04 | 0.08 | -0.12 - 0.21 | 0.588 | 0.647 | 0.01 |
| | | CBCL - Rule Breaking | 0.21 | 0.06 | 0.09 - 0.33 | <0.001 | **0.002** | 0.23 |
| | | CBCL - Aggressive | 0.16 | 0.07 | 0.02 - 0.30 | 0.025 | **0.042** | 0.09 |
| | | PQB - Distress Score | 0.23 | 0.13 | -0.02 - 0.48 | 0.068 | 0.100 | 0.06 |
| African | Family History | NIH Total Cognitive Score | 1.04 | 2.25 | -3.38 - 5.46 | 0.644 | 0.827 | 0.01 |
| | | CBCL - Total | 7.91 | 1.72 | 4.54 - 11.27 | <0.001 | **<0.001** | 1.24 |
| | | CBCL - Anxious Depressed | 3.07 | 0.77 | 1.57 - 4.58 | <0.001 | **<0.001** | 0.94 |
| | | CBCL - Withdrawn | 2.42 | 0.86 | 0.73 - 4.11 | 0.005 | **0.016** | 0.46 |
| | | CBC - Somatic | 2.86 | 0.86 | 1.17 - 4.54 | <0.001 | **0.003** | 0.65 |
| | | CBCL - Social | 4.10 | 0.72 | 2.70 - 5.51 | <0.001 | **<0.001** | 1.89 |
| | | CBCL - Thought | 4.03 | 0.88 | 2.30 - 5.76 | <0.001 | **<0.001** | 1.22 |
| | | CBCL - Attention | 1.82 | 0.98 | -0.11 - 3.75 | 0.064 | 0.176 | 0.20 |

| | | | | | | | | |
|---|---|---|---|---|---|---|---|---|
| | | CBCL - Rule Breaking | 1.43 | 0.87 | -0.28 - 3.15 | 0.101 | 0.246 | 0.16 |
| | | CBCL - Aggressive | 3.05 | 0.92 | 1.26 - 4.85 | <0.001 | **0.003** | 0.65 |
| | | PQB - Distress Score | 2.47 | 1.77 | -1.01 - 5.94 | 0.164 | 0.328 | 0.11 |
| | SCZ PRS | NIH Total Cognitive Score | 0.19 | 0.45 | -0.69 - 1.07 | 0.677 | 0.827 | 0.01 |
| | | CBCL - Total | -0.09 | 0.36 | -0.79 - 0.62 | 0.812 | 0.940 | 0.00 |
| | | CBCL - Anxious Depressed | -0.02 | 0.16 | -0.33 - 0.30 | 0.924 | 0.988 | 0.00 |
| | | CBCL - Withdrawn | -0.29 | 0.18 | -0.64 - 0.07 | 0.114 | 0.250 | 0.15 |
| | | CBC - Somatic | 0.00 | 0.18 | -0.35 - 0.35 | 0.988 | 0.988 | 0.00 |
| | | CBCL - Social | 0.09 | 0.15 | -0.21 - 0.38 | 0.571 | 0.827 | 0.02 |
| | | CBCL - Thought | 0.01 | 0.18 | -0.35 - 0.37 | 0.973 | 0.988 | 0.00 |
| | | CBCL - Attention | 0.09 | 0.20 | -0.30 - 0.49 | 0.644 | 0.827 | 0.01 |
| | | CBCL - Rule Breaking | -0.13 | 0.18 | -0.49 - 0.22 | 0.458 | 0.725 | 0.03 |
| | | CBCL - Aggressive | -0.17 | 0.19 | -0.54 - 0.21 | 0.381 | 0.699 | 0.05 |
| | | PQB - Distress Score | 0.28 | 0.37 | -0.46 - 1.01 | 0.461 | 0.725 | 0.03 |
| American Admixed | Family History | NIH Total Cognitive Score | 4.28 | 3.68 | -2.93 - 11.48 | 0.245 | 0.337 | 0.11 |
| | | CBCL - Total | 6.40 | 2.60 | 1.30 - 11.49 | 0.014 | **0.044** | 0.49 |
| | | CBCL - Anxious Depressed | 3.93 | 1.38 | 1.22 - 6.64 | 0.004 | **0.024** | 0.66 |
| | | CBCL - Withdrawn | 3.57 | 1.45 | 0.73 - 6.42 | 0.014 | **0.044** | 0.49 |
| | | CBC - Somatic | 3.26 | 1.50 | 0.33 - 6.20 | 0.029 | 0.075 | 0.39 |
| | | CBCL - Social | 4.16 | 1.14 | 1.93 - 6.39 | 0.000 | **0.006** | 1.08 |
| | | CBCL - Thought | 3.42 | 1.35 | 0.77 - 6.08 | 0.011 | **0.044** | 0.52 |
| | | CBCL - Attention | 4.70 | 1.45 | 1.87 - 7.54 | 0.001 | **0.008** | 0.86 |
| | | CBCL - Rule Breaking | 1.96 | 1.06 | -0.11 - 4.04 | 0.064 | 0.106 | 0.28 |
| | | CBCL - Aggressive | 4.11 | 1.26 | 1.64 - 6.58 | 0.001 | **0.008** | 0.86 |
| | | PQB - Distress Score | 5.39 | 2.68 | 0.14 - 10.63 | 0.044 | 0.084 | 0.33 |
| | SCZ PRS | NIH Total Cognitive Score | -0.20 | 0.47 | -1.12 - 0.72 | 0.670 | 0.737 | 0.02 |
| | | CBCL - Total | 0.67 | 0.33 | 0.01 - 1.32 | 0.046 | 0.084 | 0.33 |
| | | CBCL - Anxious Depressed | 0.28 | 0.18 | -0.07 - 0.63 | 0.119 | 0.174 | 0.20 |
| | | CBCL - Withdrawn | 0.37 | 0.19 | 0.01 - 0.74 | 0.045 | 0.084 | 0.33 |
| | | CBC - Somatic | 0.12 | 0.19 | -0.26 - 0.49 | 0.539 | 0.624 | 0.03 |
| | | CBCL - Social | 0.31 | 0.14 | -0.03 - 0.60 | 0.031 | 0.075 | 0.38 |
| | | CBCL - Thought | 0.17 | 0.17 | -0.17 - 0.51 | 0.337 | 0.435 | 0.08 |
| | | CBCL - Attention | 0.02 | 0.19 | -0.35 - 0.38 | 0.921 | 0.921 | 0.00 |
| | | CBCL - Rule Breaking | 0.25 | 0.14 | -0.02 - 0.52 | 0.067 | 0.106 | 0.27 |
| | | CBCL - Aggressive | 0.12 | 0.16 | -0.20 - 0.44 | 0.451 | 0.551 | 0.05 |
| | | PQB - Distress Score | 0.07 | 0.34 | -0.60 - 0.75 | 0.829 | 0.869 | 0.00 |

Table S3. Independent associations of schizophrenia polygenic risk scores (SCZ-PRS) and family history of psychosis (SCZ-FH) with diagnostic phenotypes derived from the Kiddie Schedule for Affective Disorders and Schizophrenia (KSADS) or Child Behavior Checklist (CBCL), including income-to-needs as a covariate. FDR P refers to the false discovery rate–corrected p value; PVE refers to the percentage of variance explained.

| **Ancestry** | **Genetic Risk Measure** | **Phenotype** | **OR** | **SE** | **95% CI** | **Nominal P** | **FDR P** | **PVE (%)** |
|---|---|---|---|---|---|---|---|---|
| All | Family History | KSADS - ADHD | 2.24 | 0.21 | 1.50 - 3.35 | <0.001 | **<0.001** | 0.22 |
| | | CBCL - ADHD | 2.95 | 0.26 | 1.78 - 4.90 | <0.001 | **<0.001** | 0.28 |
| | | KSADS - Anxiety Disorders | 4.09 | 0.21 | 2.69 - 6.22 | <0.001 | **<0.001** | 0.69 |
| | | KSADS - Anxiety disorders w/ PTSD | 4.25 | 0.21 | 2.81 - 6.43 | <0.001 | **<0.001** | 0.74 |
| | | KSADS - Depressive Disorders | 2.69 | 0.28 | 1.55 - 4.66 | <0.001 | **0.001** | 0.19 |
| | | KSADS - Conduct Disorder | 1.95 | 0.23 | 1.25 - 3.05 | 0.003 | **0.006** | 0.13 |
| | SCZ PRS | KSADS - ADHD | 1.04 | 0.03 | 0.98 - 1.09 | 0.209 | 0.279 | 0.02 |
| | | CBCL - ADHD | 0.95 | 0.04 | 0.88 - 1.04 | 0.276 | 0.331 | 0.02 |
| | | KSADS - Anxiety Disorders | 1.05 | 0.03 | 0.98 - 1.11 | 0.152 | 0.229 | 0.03 |
| | | KSADS - Anxiety disorders w/ PTSD | 1.05 | 0.03 | 0.98 - 1.11 | 0.150 | 0.229 | 0.03 |
| | | KSADS - Depressive Disorders | 1.04 | 0.05 | 0.94 - 1.15 | 0.422 | 0.422 | 0.01 |
| | | KSADS - Conduct Disorder | 1.03 | 0.03 | 0.97 - 1.09 | 0.360 | 0.393 | 0.01 |
| European | Family History | KSADS - ADHD | 2.46 | 0.33 | 1.29 - 4.68 | 0.006 | **0.019** | 0.16 |
| | | CBCL - ADHD | 4.86 | 0.42 | 2.14 - 11.04 | <0.001 | **<0.001** | 0.35 |
| | | KSADS - Anxiety Disorders | 3.68 | 0.33 | 1.93 - 7.05 | <0.001 | **<0.001** | 0.36 |
| | | KSADS - Anxiety disorders w/ PTSD | 3.67 | 0.33 | 1.92 - 7.00 | <0.001 | **<0.001** | 0.36 |
| | | KSADS - Depressive Disorders | 2.42 | 0.55 | 0.82 - 7.18 | 0.110 | 0.147 | 0.06 |
| | | KSADS - Conduct Disorder | 1.36 | 0.37 | 0.65 - 2.81 | 0.413 | 0.450 | 0.01 |
| | SCZ PRS | KSADS - ADHD | 1.07 | 0.04 | 0.99 - 1.15 | 0.062 | 0.124 | 0.08 |
| | | CBCL - ADHD | 1.01 | 0.06 | 0.91 - 1.13 | 0.802 | 0.802 | 0.00 |
| | | KSADS - Anxiety Disorders | 1.07 | 0.04 | 0.99 - 1.15 | 0.085 | 0.145 | 0.07 |
| | | KSADS - Anxiety disorders w/ PTSD | 1.08 | 0.04 | 1.00 - 1.16 | 0.049 | 0.118 | 0.09 |
| | | KSADS - Depressive Disorders | 1.11 | 0.07 | 0.97 - 1.27 | 0.131 | 0.157 | 0.06 |
| | | KSADS - Conduct Disorder | 1.07 | 0.04 | 0.99 - 1.15 | 0.096 | 0.145 | 0.06 |
| African | Family History | KSADS - ADHD | 1.86 | 0.31 | 1.01 - 3.43 | 0.047 | 0.114 | 0.27 |
| | | CBCL - ADHD | 1.74 | 0.41 | 0.78 - 3.84 | 0.173 | 0.347 | 0.15 |
| | | KSADS - Anxiety Disorders | 4.26 | 0.33 | 2.23 - 8.12 | <0.001 | **<0.001** | 1.62 |
| | | KSADS - Anxiety disorders w/ PTSD | 4.79 | 0.32 | 2.54 - 9.06 | <0.001 | **<0.001** | 1.92 |
| | | KSADS - Depressive Disorders | 3.18 | 0.38 | 1.52 - 6.67 | 0.002 | **0.009** | 0.73 |
| | | KSADS - Conduct Disorder | 2.18 | 0.33 | 1.13 - 4.17 | 0.019 | 0.058 | 0.41 |
| | SCZ PRS | KSADS - ADHD | 1.05 | 0.07 | 0.91 - 1.21 | 0.502 | 0.687 | 0.03 |
| | | CBCL - ADHD | 0.95 | 0.10 | 0.78 - 1.16 | 0.644 | 0.749 | 0.02 |
| | | KSADS - Anxiety Disorders | 1.04 | 0.09 | 0.87 - 1.23 | 0.687 | 0.749 | 0.01 |
| | | KSADS - Anxiety disorders w/ PTSD | 1.01 | 0.09 | 0.86 - 1.20 | 0.891 | 0.891 | 0.00 |
| | | KSADS - Depressive Disorders | 0.93 | 0.11 | 0.75 - 1.15 | 0.502 | 0.687 | 0.03 |
| | | KSADS - Conduct Disorder | 1.06 | 0.08 | 0.90 - 1.24 | 0.515 | 0.687 | 0.03 |
| American Admixed | Family History | KSADS - ADHD | 3.66 | 0.54 | 1.28 - 10.50 | 0.016 | **0.038** | 0.58 |
| | | CBCL - ADHD | 6.02 | 0.66 | 1.65 - 21.94 | 0.006 | **0.029** | 0.84 |
| | | KSADS - Anxiety Disorders | 5.21 | 0.57 | 1.70 - 15.99 | 0.004 | **0.029** | 0.91 |
| | | KSADS - Anxiety disorders w/ PTSD | 4.66 | 0.57 | 1.52 - 14.31 | 0.007 | **0.029** | 0.78 |
| | | KSADS - Depressive Disorders | 2.64 | 0.75 | 0.60 - 11.55 | 0.199 | 0.299 | 0.15 |
| | | KSADS - Conduct Disorder | 4.17 | 0.63 | 1.22 - 14.23 | 0.022 | **0.045** | 0.53 |
| | SCZ PRS | KSADS - ADHD | 0.97 | 0.08 | 0.83 - 1.14 | 0.740 | 0.808 | 0.01 |
| | | CBCL - ADHD | 0.90 | 0.12 | 0.71 - 1.13 | 0.362 | 0.434 | 0.09 |
| | | KSADS - Anxiety Disorders | 1.13 | 0.10 | 0.94 - 1.37 | 0.200 | 0.299 | 0.17 |
| | | KSADS - Anxiety disorders w/ PTSD | 1.10 | 0.10 | 0.91 - 1.32 | 0.331 | 0.434 | 0.10 |
| | | KSADS - Depressive Disorders | 1.38 | 0.13 | 1.07 - 1.77 | 0.012 | **0.036** | 0.56 |
| | | KSADS - Conduct Disorder | 1.02 | 0.10 | 0.83 - 1.25 | 0.874 | 0.874 | 0.00 |

Table S4. Joint model associations of schizophrenia polygenic risk scores (SCZ-PRS) and family history of psychosis (SCZ-FH) with total cognitive score derived from the NIH-Toolbox, Child Behavior Checklist (CBCL) scores, and prodromal questionnaire (PQB) scores, including income-to-needs as a covariate. FDR P refers to the false discovery rate–corrected p value; PVE refers to the percentage of variance explained.

| **Ancestry** | **Phenotype** | **Family History of Psychosis (Joint Model)** | | | | | | **SCZ PRS (Joint Model)** | | | | | | **Joint PVE** |
|---|---|---|---|---|---|---|---|---|---|---|---|---|---|---|
| | | *B* | *SE* | *95% CI* | *Nominal P* | *FDR P* | *PVE* | *B* | *SE* | *95% CI* | *Nominal P* | *FDR P* | *PVE* | |
| All | NIH Total Cognitive Score | 0.92 | 1.49 | -1.99 - 3.83 | 0.536 | 0.610 | 0.00 | -0.48 | 0.18 | -0.83 - -0.13 | 0.007 | **0.013** | 0.09 | 0.10 |
| | CBCL - Total | 7.17 | 1.02 | 5.17 - 9.16 | <0.001 | **<0.001** | 0.60 | 0.15 | 0.12 | -0.09 - 0.39 | 0.223 | 0.289 | 0.02 | 0.62 |
| | CBCL - Anxious Depressed | 2.77 | 0.54 | 1.71 - 3.83 | <0.001 | **<0.001** | 0.31 | 0.09 | 0.07 | -0.04 - 0.22 | 0.155 | 0.227 | 0.02 | 0.34 |
| | CBCL - Withdrawn | 2.10 | 0.52 | 1.08 - 3.12 | <0.001 | **<0.001** | 0.19 | 0.03 | 0.06 | -0.10 - 0.15 | 0.672 | 0.704 | 0.00 | 0.20 |
| | CBC - Somatic | 2.82 | 0.55 | 1.74 - 3.91 | <0.001 | **<0.001** | 0.31 | -0.09 | 0.07 | -0.23 - 0.04 | 0.165 | 0.227 | 0.02 | 0.33 |
| | CBCL - Social | 3.58 | 0.42 | 2.75 - 4.41 | <0.001 | **<0.001** | 0.85 | 0.03 | 0.05 | -0.07 - 0.13 | 0.528 | 0.610 | 0.00 | 0.86 |
| | CBCL - Thought | 4.31 | 0.55 | 3.24 - 5.39 | <0.001 | **<0.001** | 0.74 | 0.09 | 0.07 | -0.04 - 0.22 | 0.165 | 0.227 | 0.02 | 0.77 |
| | CBCL - Attention | 3.30 | 0.57 | 2.18 - 4.42 | <0.001 | **<0.001** | 0.40 | -0.01 | 0.07 | -0.15 - 0.12 | 0.846 | 0.846 | 0.00 | 0.40 |
| | CBCL - Rule Breaking | 1.89 | 0.44 | 1.02 - 2.75 | <0.001 | **<0.001** | 0.22 | 0.11 | 0.05 | 0.01 - 0.22 | 0.036 | 0.066 | 0.05 | 0.28 |
| | CBCL - Aggressive | 3.18 | 0.51 | 2.18 - 4.17 | <0.001 | **<0.001** | 0.47 | 0.04 | 0.06 | -0.08 - 0.16 | 0.555 | 0.610 | 0.00 | 0.48 |
| | PQB - Distress Score | 3.22 | 0.94 | 1.37 - 5.06 | 0.001 | **0.001** | 0.14 | 0.20 | 0.12 | -0.03 - 0.42 | 0.090 | 0.152 | 0.03 | 0.18 |
| European | NIH Total Cognitive Score | -0.50 | 2.31 | -5.02 - 4.02 | 0.828 | 0.867 | 0.00 | -0.68 | 0.22 | -1.12 - -0.24 | 0.002 | **0.006** | 0.18 | 0.18 |
| | CBCL - Total | 6.27 | 1.53 | 3.27 - 9.26 | <0.001 | **<0.001** | 0.31 | 0.30 | 0.15 | 0.01 - 0.59 | 0.045 | 0.070 | 0.08 | 0.40 |
| | CBCL - Anxious Depressed | 1.88 | 0.87 | 0.18 - 3.58 | 0.030 | 0.055 | 0.09 | 0.19 | 0.08 | 0.03 - 0.36 | 0.021 | **0.043** | 0.10 | 0.19 |
| | CBCL - Withdrawn | 1.05 | 0.78 | -0.48 - 2.57 | 0.178 | 0.230 | 0.03 | 0.09 | 0.08 | -0.06 - 0.24 | 0.224 | 0.273 | 0.03 | 0.06 |
| | CBC - Somatic | 2.67 | 0.85 | 1.00 - 4.34 | 0.002 | **0.005** | 0.18 | -0.10 | 0.08 | -0.26 - 0.07 | 0.251 | 0.291 | 0.02 | 0.20 |
| | CBCL - Social | 2.71 | 0.63 | 1.48 - 3.95 | <0.001 | **<0.001** | 0.34 | 0.00 | 0.06 | -0.12 - 0.12 | 0.967 | 0.967 | 0.00 | 0.34 |
| | CBCL - Thought | 4.84 | 0.85 | 3.18 - 6.50 | <0.001 | **<0.001** | 0.61 | 0.12 | 0.08 | -0.04 - 0.29 | 0.130 | 0.179 | 0.04 | 0.66 |
| | CBCL - Attention | 4.31 | 0.85 | 2.63 - 5.98 | <0.001 | **<0.001** | 0.47 | 0.03 | 0.08 | -0.13 - 0.19 | 0.724 | 0.796 | 0.00 | 0.48 |
| | CBCL - Rule Breaking | 2.18 | 0.62 | 0.96 - 3.40 | <0.001 | **0.002** | 0.23 | 0.20 | 0.06 | 0.08 - 0.32 | <0.001 | **0.003** | 0.21 | 0.46 |
| | CBCL - Aggressive | 2.82 | 0.74 | 1.36 - 4.27 | <0.001 | **<0.001** | 0.27 | 0.15 | 0.07 | 0.01 - 0.29 | 0.035 | 0.060 | 0.08 | 0.36 |
| | PQB - Distress Score | 3.42 | 1.30 | 0.86 - 5.97 | 0.009 | **0.019** | 0.13 | 0.22 | 0.13 | -0.03 - 0.47 | 0.085 | 0.125 | 0.06 | 0.19 |
| African | NIH Total Cognitive Score | 1.01 | 2.25 | -3.41 - 5.43 | 0.653 | 0.840 | 0.01 | 0.18 | 0.45 | -0.70 - 1.06 | 0.687 | 0.840 | 0.01 | 0.02 |
| | CBCL - Total | 7.92 | 1.72 | 4.55 - 11.29 | <0.001 | **<0.001** | 1.24 | -0.12 | 0.36 | -0.81 - 0.58 | 0.742 | 0.859 | 0.01 | 1.24 |
| | CBCL - Anxious Depressed | 3.07 | 0.77 | 1.57 - 4.58 | <0.001 | **<0.001** | 0.94 | -0.03 | 0.16 | -0.34 - 0.29 | 0.865 | 0.952 | 0.00 | 0.94 |
| | CBCL - Withdrawn | 2.44 | 0.86 | 0.75 - 4.13 | 0.005 | **0.014** | 0.47 | -0.30 | 0.18 | -0.65 - 0.06 | 0.102 | 0.224 | 0.16 | 0.62 |
| | CBC - Somatic | 2.86 | 0.86 | 1.17 - 4.54 | <0.001 | **0.003** | 0.65 | -0.01 | 0.18 | -0.36 - 0.33 | 0.936 | 0.957 | 0.00 | 0.65 |
| | CBCL - Social | 4.10 | 0.72 | 2.69 - 5.51 | <0.001 | **<0.001** | 1.89 | 0.07 | 0.15 | -0.22 - 0.36 | 0.645 | 0.840 | 0.01 | 1.91 |
| | CBCL - Thought | 4.03 | 0.88 | 2.30 - 5.76 | <0.001 | **<0.001** | 1.22 | -0.01 | 0.18 | -0.37 - 0.35 | 0.957 | 0.957 | 0.00 | 1.22 |
| | CBCL - Attention | 1.81 | 0.98 | -0.12 - 3.74 | 0.065 | 0.180 | 0.20 | 0.09 | 0.20 | -0.31 - 0.48 | 0.670 | 0.840 | 0.01 | 0.21 |
| | CBCL - Rule Breaking | 1.45 | 0.87 | -0.26 - 3.16 | 0.098 | 0.224 | 0.16 | -0.14 | 0.18 | -0.49 - 0.21 | 0.439 | 0.742 | 0.04 | 0.19 |
| | CBCL - Aggressive | 3.07 | 0.92 | 1.27 - 4.87 | <0.001 | **0.003** | 0.66 | -0.18 | 0.19 | -0.55 - 0.19 | 0.345 | 0.632 | 0.05 | 0.70 |
| | PQB - Distress Score | 2.44 | 1.77 | -1.03 - 5.92 | 0.168 | 0.098 | 0.11 | 0.27 | 0.37 | -0.47 - 1.00 | 0.476 | 0.913 | 0.03 | 0.14 |
| American Admixed | NIH Total Cognitive Score | 4.32 | 3.68 | -2.90 - 11.53 | 0.241 | 0.331 | 0.12 | -0.21 | 0.47 | -1.14 - 0.71 | 0.650 | 0.715 | 0.02 | 0.13 |
| | CBCL - Total | 6.26 | 2.60 | 1.17 - 11.35 | 0.016 | 0.050 | 0.47 | 0.64 | 0.33 | -0.01 - 1.30 | 0.053 | 0.098 | 0.30 | 0.80 |
| | CBCL - Anxious Depressed | 3.87 | 1.38 | 1.17 - 6.58 | 0.005 | **0.028** | 0.64 | 0.26 | 0.18 | -0.08 - 0.61 | 0.137 | 0.200 | 0.18 | 0.84 |
| | CBCL - Withdrawn | 3.50 | 1.45 | 0.65 - 6.34 | 0.016 | 0.050 | 0.47 | 0.36 | 0.19 | -0.00 - 0.73 | 0.052 | 0.098 | 0.31 | 0.80 |
| | CBC - Somatic | 3.24 | 1.50 | 0.30 - 6.18 | 0.031 | 0.084 | 0.38 | 0.11 | 0.19 | -0.27 - 0.48 | 0.579 | 0.670 | 0.03 | 0.41 |
| | CBCL - Social | 4.09 | 1.14 | 1.86 - 6.32 | 0.000 | **0.007** | 1.05 | 0.30 | 0.14 | 0.01 - 0.58 | 0.039 | 0.096 | 0.34 | 1.43 |
| | CBCL - Thought | 3.39 | 1.35 | 0.74 - 6.04 | 0.012 | 0.050 | 0.51 | 0.15 | 0.17 | -0.18 - 0.49 | 0.373 | 0.482 | 0.06 | 0.59 |
| | CBCL - Attention | 4.70 | 1.45 | 1.86 - 7.54 | 0.001 | **0.009** | 0.86 | 0.00 | 0.18 | -0.36 - 0.36 | 0.994 | 0.994 | 0.00 | 0.86 |
| | CBCL - Rule Breaking | 1.91 | 1.06 | -0.16 - 3.98 | 0.071 | 0.118 | 0.27 | 0.24 | 0.14 | -0.02 - 0.51 | 0.075 | 0.118 | 0.26 | 0.54 |
| | CBCL - Aggressive | 4.09 | 1.26 | 1.62 - 6.56 | 0.001 | **0.009** | 0.85 | 0.11 | 0.16 | -0.21 - 0.43 | 0.501 | 0.612 | 0.04 | 0.90 |
| | PQB - Distress Score | 5.37 | 2.68 | 0.13 - 10.62 | 0.045 | 0.336 | 0.33 | 0.06 | 0.34 | -0.62 - 0.73 | 0.871 | 0.748 | 0.00 | 0.33 |

Table S5. Joint model associations of schizophrenia polygenic risk scores (SCZ-PRS) and family history of psychosis (SCZ-FH) with diagnostic phenotypes derived from the Kiddie Schedule for Affective Disorders and Schizophrenia (KSADS) or Child Behavior Checklist (CBCL), including income-to-needs as a covariate. FDR P refers to the false discovery rate–corrected p value; PVE refers to the percentage of variance explained.

| **Ancestry** | **Phenotype** | **Family History of Psychosis (Joint Model)** | | | | | | **SCZ PRS (Joint Model)** | | | | | | **Joint PVE** |
|---|---|---|---|---|---|---|---|---|---|---|---|---|---|---|
| | | *OR* | *SE* | *95% CI* | *Nominal P* | *FDR P* | *PVE* | *OR* | *SE* | *95% CI* | *Nominal P* | *FDR P* | *PVE* | |
| All | KSADS - ADHD | 2.23 | 0.21 | 1.49 - 3.34 | <0.001 | **<0.001** | 0.22 | 1.03 | 0.03 | 0.98 - 1.09 | 0.240 | 0.302 | 0.02 | 0.24 |
| | CBCL - ADHD | 2.97 | 0.26 | 1.79 - 4.93 | <0.001 | **<0.001** | 0.28 | 0.95 | 0.04 | 0.87 - 1.04 | 0.252 | 0.302 | 0.02 | 0.30 |
| | KSADS - Anxiety Disorders | 4.07 | 0.21 | 2.67 - 6.18 | <0.001 | **<0.001** | 0.69 | 1.04 | 0.03 | 0.98 - 1.11 | 0.198 | 0.298 | 0.02 | 0.89 |
| | KSADS - Anxiety disorders w/ PTSD | 4.22 | 0.21 | 2.79 - 6.38 | <0.001 | **<0.001** | 0.74 | 1.04 | 0.03 | 0.98 - 1.11 | 0.197 | 0.298 | 0.02 | 0.77 |
| | KSADS - Depressive Disorders | 2.68 | 0.28 | 1.54 - 4.64 | <0.001 | **0.001** | 0.19 | 1.04 | 0.05 | 0.94 - 1.15 | 0.450 | 0.450 | 0.01 | 0.19 |
| | KSADS - Conduct Disorder | 1.95 | 0.23 | 1.25 - 3.04 | 0.003 | **0.007** | 0.13 | 1.03 | 0.03 | 0.97 - 1.09 | 0.396 | 0.432 | 0.01 | 0.14 |
| European | KSADS - ADHD | 2.40 | 0.33 | 1.26 - 4.58 | 0.008 | **0.024** | 0.16 | 1.06 | 0.04 | 0.99 - 1.14 | 0.079 | 0.157 | 0.07 | 0.23 |
| | CBCL - ADHD | 4.86 | 0.42 | 2.14 - 11.03 | <0.001 | **<0.001** | 0.35 | 1.01 | 0.06 | 0.90 - 1.13 | 0.923 | 0.923 | 0.00 | 0.35 |
| | KSADS - Anxiety Disorders | 3.61 | 0.33 | 1.88 - 6.91 | <0.001 | **<0.001** | 0.35 | 1.06 | 0.04 | 0.99 - 1.14 | 0.117 | 0.167 | 0.05 | 0.42 |
| | KSADS - Anxiety disorders w/ PTSD | 3.58 | 0.33 | 1.87 - 6.83 | <0.001 | **<0.001** | 0.35 | 1.07 | 0.04 | 0.99 - 1.15 | 0.070 | 0.157 | 0.07 | 0.44 |
| | KSADS - Depressive Disorders | 2.35 | 0.56 | 0.79 - 7.00 | 0.125 | 0.167 | 0.06 | 1.11 | 0.07 | 0.97 - 1.27 | 0.146 | 0.175 | 0.05 | 0.11 |
| | KSADS - Conduct Disorder | 1.32 | 0.37 | 0.64 - 2.74 | 0.454 | 0.495 | 0.01 | 1.06 | 0.04 | 0.99 - 1.15 | 0.103 | 0.167 | 0.06 | 0.07 |
| African | KSADS - ADHD | 1.85 | 0.31 | 1.00 - 3.41 | 0.049 | 0.117 | 0.26 | 1.05 | 0.07 | 0.91 - 1.21 | 0.520 | 0.714 | 0.03 | 0.29 |
| | CBCL - ADHD | 1.74 | 0.41 | 0.79 - 3.86 | 0.172 | 0.344 | 0.15 | 0.95 | 0.10 | 0.78 - 1.16 | 0.635 | 0.762 | 0.02 | 0.16 |
| | KSADS - Anxiety Disorders | 4.24 | 0.33 | 2.22 - 8.10 | <0.001 | **<0.001** | 1.61 | 1.03 | 0.09 | 0.86 - 1.22 | 0.753 | 0.821 | 0.01 | 1.62 |
| | KSADS - Anxiety disorders w/ PTSD | 4.79 | 0.32 | 2.54 - 9.06 | <0.001 | **<0.001** | 1.92 | 1.00 | 0.09 | 0.85 - 1.19 | 0.979 | 0.979 | 0.00 | 1.92 |
| | KSADS - Depressive Disorders | 3.21 | 0.38 | 1.53 - 6.74 | 0.002 | **0.008** | 0.73 | 0.92 | 0.11 | 0.74 - 1.14 | 0.460 | 0.714 | 0.04 | 0.76 |
| | KSADS - Conduct Disorder | 2.17 | 0.33 | 1.13 - 4.16 | 0.020 | 0.060 | 0.41 | 1.05 | 0.08 | 0.90 - 1.24 | 0.535 | 0.714 | 0.03 | 0.44 |
| American Admixed | KSADS - ADHD | 3.68 | 0.54 | 1.29 - 10.56 | 0.015 | **0.036** | 0.59 | 0.97 | 0.08 | 0.83 - 1.13 | 0.683 | 0.745 | 0.02 | 0.60 |
| | CBCL - ADHD | 6.14 | 0.66 | 1.69 - 22.26 | 0.006 | **0.031** | 0.86 | 0.89 | 0.12 | 0.70 - 1.12 | 0.324 | 0.425 | 0.11 | 0.95 |
| | KSADS - Anxiety Disorders | 5.19 | 0.57 | 1.68 - 15.99 | 0.004 | **0.031** | 0.89 | 1.13 | 0.10 | 0.93 - 1.36 | 0.213 | 0.320 | 0.15 | 1.06 |
| | KSADS - Anxiety disorders w/ PTSD | 4.63 | 0.57 | 1.50 - 14.29 | 0.008 | **0.031** | 0.77 | 1.09 | 0.09 | 0.91 - 1.31 | 0.354 | 0.425 | 0.08 | 0.86 |
| | KSADS - Depressive Disorders | 2.60 | 0.77 | 0.58 - 11.75 | 0.213 | 0.320 | 0.15 | 1.37 | 0.13 | 1.07 - 1.76 | 0.012 | **0.036** | 0.56 | 0.70 |
| | KSADS - Conduct Disorder | 4.16 | 0.63 | 1.22 - 14.23 | 0.023 | **0.046** | 0.52 | 1.01 | 0.10 | 0.82 - 1.23 | 0.944 | 0.944 | 0.00 | 0.53 |
